# Remote exposure to secondhand tobacco smoke is associated with lower exercise capacity through effects on oxygen pulse, a proxy of cardiac stroke volume

**DOI:** 10.1101/2021.05.29.21258054

**Authors:** Siyang Zeng, Michelle Dunn, Warren M Gold, Jorge Kizer, Mehrdad Arjomandi

**Affiliations:** Department of Biomedical Informatics and Medical Education, University of Washington, Seattle, WA, USA; Pulmonary and Critical Care Section, San Francisco Veterans Affairs Medical Center, San Francisco, California, USA; Department of Medicine, University of California, San Francisco, California, USA; Cardiology Section, San Francisco Veterans Affairs Medical Center, San Francisco, California, USA; Department of Epidemiology and Biostatistics, University of California, San Francisco, California, USA

**Keywords:** Exercise capacity, Secondhand smoke, Cardiac stroke volume, Oxygen-pulse, Air trapping

## Abstract

**Background:** Past exposure to secondhand tobacco smoke (SHS) is associated with exercise limitation. Pulmonary factors including air trapping contribute to this limitation but the contribution of cardiovascular factors is unclear.

**Objective:** To determine contribution of cardiovascular mechanisms to SHS-associated exercise limitation.

**Methods:** We examined the cardiovascular responses to maximum effort exercise in 245 never-smokers with remote, prolonged occupational exposure to SHS and no known history of cardiovascular disease. We estimated the contribution of oxygen-pulse (proxy for cardiac stroke volume) and changes in systolic (SBP) and diastolic blood pressures (DBP) and heart rate (HR) towards exercise capacity, and examined whether the association of SHS with exercise capacity was mediated through these variables.

**Results:** At peak exercise (highest workload completed [Watts_Peak_]=156±46 watts [135±33 %predicted]), oxygen consumption (VO_2Peak_) and oxygen-pulse (O_2_-Pulse_Peak_) were 1,557±476 mL/min (100±24 %predicted) and 11.0±3.0 mL/beat (116±25 %predicted), respectively, with 29% and 3% participants not achieving their predicted normal range. Oxygen saturation at peak exercise was 98±1% and remained >93% in all participants. Sixty-six percent showed hypertensive response to exercise. In models adjusted for covariates, Watts_Peak_ was associated directly with O_2_-Pulse_Peak_, HR_Peak_, and SBP_Peak_ and inversely with SHS, air trapping (RV/TLC), and rise of SBP over workload (all P<0.01). Moreover, SHS exposure association with Watts_Peak_ was substantially (41%) mediated through its effect on O_2_-Pulse_Peak_ (P<0.038). Although not statistically significant, a considerable proportion (36%) of air trapping effect on Watts_Peak_ seemed to be mediated through O_2_-Pulse_Peak_ (P=0.078). The likelihood of having baseline respiratory symptoms (modified Medical Research Council score ≥1) was associated with steeper rise in SBP over workload (P<0.01).

**Conclusion:** In a never-smoker population with remote exposure to SHS, abnormal escalation of afterload and an SHS-associated reduction in cardiac output contributed to lower exercise capacity.

**Key messages:** *What is the key question?:* What are the cardiovascular health effects of past exposure to secondhand tobacco smoke in never-smokers? How do pulmonary and cardiovascular systems interact in this setting?

*What is the bottom line?:* Healthy never-smokers with history of remote past exposure to secondhand tobacco smoke have an abnormal cardiovascular response to exercise, which is characterized by a stroke volume and thus an exercise capacity that are reduced proportional to their years of exposure to secondhand tobacco smoke.

*Why read on?:* The abnormal cardiovascular response to exercise in this population reveals the presence of an occult or subclinical pathology that impairs the cardiopulmonary functional reserve and reduces the efficiency of body’s oxygen delivery machinery, which could be disadvantageous during the times of increased cardiopulmonary output demands as in physiological distress or disease.

## INTRODUCTION

Secondhand tobacco smoke (SHS) remains a major public health problem.^1, 2^ Although SHS exposure among nonsmokers in the United States has declined from 88% in 1988 to 25% in 2014, the rate of decline has plateaued with one in four nonsmokers, including 14 million children, continuing to be exposed to SHS annually between 2011 and 2014.^3^ This substantial continued exposure is important particularly as the generations that endured the highest amount of SHS exposure are aging, a process which could accentuate the SHS-related health problems that may have been previously too subtle to be recognized.

Although immediate health effects of exposure to SHS have been studied,^4–9^ its long-term consequences, particularly the effect of remote exposures, have been more difficult to examine in part due to challenges with exposure assessment. Never-smoking flight crews who worked on commercial aircrafts before the enactment of the smoking ban were exposed to heavy SHS in aircraft cabin for many years, in a range similar to the nicotine exposure burden experienced by “light” smokers.^10, 11^ The regularity of this intense exposure in the cabin work environment lends itself to relatively accurate SHS exposure quantification through employment history,^12^ and makes the exposed flight crew a unique population in which the long-term health effects of previous exposure to SHS could be examined as a form of “natural” experiment that is also generalizable to other SHS-exposed populations.

In previous studies of never-smoking flight crews with a history of remote but prolonged exposure to SHS in aircraft cabin, we examined the pulmonary health effects of long-term exposure to SHS and showed the association of those outcomes with the number of years during which the flight crew were exposed to SHS in the aircraft cabin.^13–15^ While this never-smoking SHS-exposed cohort had no evidence of spirometric chronic obstructive pulmonary disease (COPD) (a preserved ratio of forced expiratory volume in 1 second to forced vital capacity [FEV_1_/FVC]), they had abnormal lung function that were suggestive of presence of an occult early/mild obstructive lung disease.^15^ In the current study, we wished to examine the cardiovascular health effects of remote but prolonged exposure to SHS in this never-smoking cohort with evidence of early obstructive lung disease. We hypothesized that in addition to impacting pulmonary mechanisms (e.g., air trapping), prolonged exposure to SHS contributes to exercise limitation through its adverse cardiovascular health effects. To evaluate this hypothesis, we analyzed the cohort’s cardiovascular response to maximum effort cardiopulmonary exercise testing.

## METHODS

### Study Overview

This was a *post-hoc* analysis of data collected as part of the Secondhand Smoke Respiratory Health Study, an observational cohort study of nonsmoking participants with a range of occupational SHS exposure, as previously described.^13–15^ Briefly, between July 2007 and July 2015, we recruited US airline flight crewmembers with a history of occupational exposure to SHS, along with nonsmoker controls without such occupational exposure, who were participating in a larger study of cardiopulmonary health effects of prolonged remote exposure to SHS.^15^ The participants were characterized by respiratory symptom questionnaires, full pulmonary function testing (PFT), and a maximum effort cardiopulmonary exercise testing (CPET). We used the data from this cohort to perform a *post-hoc* analysis to determine the associations among exercise capacity (highest workload completed [Watts_Peak_ in Watts], volume of oxygen uptake at peak exercise [VO_2Peak_ in L/min], and cumulative work achieved [Work_Total_ in Watts-Minutes]), cardiovascular responses to maximum effort CPET (oxygen-pulse [O_2_-Pulse; a proxy for cardiac stroke volume], systolic and diastolic blood pressures [SBP and DBP], and heart rate [HR]), years of airline employment during which the participants worked in smoky cabin (cabin SHS exposure), and their interactions with each other as well as with air trapping (ratios of residual volume, or functional residual capacity, to total lung capacity; RV/TLC or FRC/TLC), which we had previously shown to be associated with exercise capacity (Watts_Peak_ and VO_2Peak_). The UCSF Institutional Review Board (IRB) approved the study protocols.

### Study Population

The Secondhand Smoke Respiratory Health Study recruited the United States (U.S.) airline flight crewmembers as part of an investigation of the potential adverse health effects of the cabin environment on those employed before and after introduction of the ban on smoking in U.S. commercial aircraft. Crewmembers were eligible to participate in the study if they had worked ≥5 years in aircraft. A referent group of “sea-level” participants who lived in San Francisco Bay area and had never been employed as airline crewmembers were also recruited. All participants were nonsmokers defined by never-smoking or, in ever smokers, a cumulative history of smoking <20 pack-years and no smoking for ≥20 years prior to enrollment. Participants with known history of pulmonary (such as asthma or COPD) or cardiovascular disease (such as coronary artery disease or heart failure) were excluded. Participants with known history of hypertension were included if their blood pressure was medically controlled as defined by SBP≤145 and DBP≤90 at the time of visit. Overall, out of the 283 participants who underwent cardiopulmonary exercise testing (CPET), 245 were included in this analysis.^15^

### Institutional Review Board Approval

The UCSF Institutional Review Board (IRB) and the San Francisco VA Medical Center Committee on Research and Development approved the study protocols. Written IRB-approved informed consent and Health Insurance Portability and Accountability Act (HIPAA) were obtained from all study participants. All participants received monetary compensation for their participation in the study.

### Patient and Public Involvement

Patients or the public were not involved in the design, conduct, reporting, or dissemination plans of our research. However, the participants did act as a referral source and referred other interested potential people to participate in our study, and in that sense participated in dissemination of the study.

### SHS Exposure Characterization

SHS exposure was characterized by a questionnaire developed by the UCSF Flight Attendant Medical Research Institute (FAMRI) Center of Excellence,^16^ and modified to acquire information on airline-related occupational history (UCSF FAMRI SHS Questionnaire), as detailed in **Supplemental Appendix** and described previously.^13, 14^

### Pulmonary Function and Cardiopulmonary Exercise Testing

Details of pulmonary function and maximum effort cardiopulmonary function testings are available in **Supplemental Appendix** and have been previously.^13, 15^

### Respiratory Symptom Scoring

Respiratory symptoms were assessed using modified Medical Research Council (mMRC) Dyspnea Scale^17^ and another self-reported questionnaire (UCSF FAMRI SHS Questionnaire) that elicited symptoms of dyspnea, cough, and participants’ perception of a decreased level of exertion compared to peers over the year preceding enrollment.^16^ A dichotomous indicator of respiratory symptoms was defined by mMRC ≥1 or report of at least one respiratory symptom on the UCSF FAMRI SHS Questionnaire. A dichotomous cause of exercise cessation (dyspnea versus fatigue or effort) was determined based on the highest Borg scale category reported by the participants at the end of the maximum effort exercise testing.

### Data Analysis

Percent predicted as well as lower and upper limits of normal (LLN and ULN) values for measures of spirometry and lung volumes at rest and cardiopulmonary responses to exercise were calculated using Global Lung Function Initiative (GLI), Quanjer et al., and Wasserman predicted formulas, repectively.^18–20^ American Thoracic Society guidelines were also used for assessment of normal ranges of cardiopulmonary exercise indices.^21^

Distributions of participants’ characteristics, pulmonary function, cardiopulmonary exercise, and SHS exposure quantification variables were examined. Changes in HR, SBP, DBP, and O_2_-Pulse with respect to the workload were approximated by estimating the slopes from linear regression modeling of the measures over workload at each stage. Peak cardiopulmonary exercise variables were estimated using the last 30-second average values obtained during the highest stage of the exercise test as described above. Cumulative work achieved throughout the exercise (Work_Total_), or the area under the curve of workload in Watts vs. time in minutes, was computed as the sum of the product of watts completed and time spent at each stage in the unit of Watts-Minute.

Associations between exercise capacity (Watts_Peak_, Work_Total_, or VO_2Peak_) and each of the cardiovascular outputs (including SBP, DBP, and O_2_-Pulse) were examined in linear regression models with adjustment for covariates (age, sex, height, and BMI unless noted otherwise). Because O_2_-Pulse (proxy for stroke volume) estimates were calculated using VO_2_ and HR, the associations and mediation analyses of O_2_-Pulse was only examined with Watts_Peak_ and Work_Total_ measures of exercise capacity and not with VO_2Peak_ to prevent bias from “mathematical coupling”.^22^

Similarly, associations between presence of respiratory symptoms (mMRC or UCSF FAMRI SHS Questionnaire) and each of the cardiovascular outputs were examined in logistic regression models with adjustment for covariates. The respective baseline variable was also adjusted whenever a slope variable was used in a model. Additionally, associations between each of the cardiovascular outputs (SBP, DBP, and O_2_-Pulse) and baseline air trapping (FRC/TLC or RV/TLC) as a pulmonary factor affecting exercise capacity and cardiovascular outputs,^23–26^ and SHS exposure were examined using linear regression models.

To assess whether associations between exercise capacity and SHS exposure or air trapping (FRC/TLC or RV/TLC) were mediated through cardiovascular outputs, we performed mediation analyses with exercise capacity (dependent variable), SHS exposure or air trapping (independent variable), and cardiovascular outputs (mediator variables), with inclusion of covariates using the “mediation” package in R.^27^ Absolute proportion of mediated effects with corresponding P values were reported.

For each analysis, the total number of participants who had complete set of data for that analysis was reported along with the results from the regression modeling or mediation analysis.

## RESULTS

### Participants’ Characteristics

The characteristics of the 245 participants included in this analysis are shown in Table 1. The participants were predominantly women (213 [87%]). All were never-smokers (<100 cigarettes lifetime) with the exception of three who had smoked ≤3 pack-years (0.7, 1, and 3 pack-years) ≥20 years prior to participation in the study. All participants had exposure to SHS with 47% and 24% reporting childhood and adult home SHS exposure, respectively. One-hundred-thirty-nine (57%) had been exposed to cabin SHS during their airline employment for an average of 16.5±9.2 years (median [IQR] <TOTAL range> of 17.1 [8.6-24.0] <0.3-35.0> years). None of the participants included in the analysis reported any history of cardiovascular or pulmonary diseases with the exception of eleven who had a history of well-controlled hypertension, nine of whom on medical management (three on lisinopril, two on atenolol, one on verapamil, one on an unknown beta-blocker, one on combination hydrochlorothiazide and benazepril, and one on an unknown medication).

**Table 1.** Participant characteristics. <u>Footnote</u>: Demographics, secondhand smoke (SHS) exposure, symptoms, and lung function in participants with preserved spirometry that underwent exercise testing. Other SHS exposure was defined as non-aircraft cabin SHS exposure outside the work or home environment such as in recreational public places. Data are presented as mean±standard deviation or number of participants with positive value for the variable (n) out of the total number of participants (N) and percentage of participants (%). Reference equations: percent predicted of normal values of spirometry, diffusing capacity, and lung volumes were calculated using Global Lung Function Initiative (GLI), Crapo, and Quanjar predicted formulas, respectively.^18–20, 47^ Abbreviations-BMI: body mass index; FEV_1_: forced expiratory volume in 1 second; FVC: forced vital capacity; FEF_25-75_: maximum airflow at mid-lung volume; FEF_75_: maximum airflow at low-lung volume; TLC: total lung capacity; RV: residual volume; FRC: functional residual capacity; DcoSB: single-breath diffusing capacity of carbon monoxide; Hgb: hemoglobin; mMRC: modified medical research council.

| **Participant characteristics** | **N=245** |
| --- | --- |
| **Demographics and anthropometrics** |  |
| Age (years) | 53.8±11.3 |
| Female sex [n (%)] | 213 (86.9%) |
| Height (cm) | 166.3±7.1 |
| Weight (kg) | 66.3±11.5 |
| BMI (kg/m<sup>2</sup>) | 23.9±3.6 |
| Hemoglobin (g/dL) | 14.2±1.3 |
| Smoking history (pack-years) [min-max] | 0-3 |
| History of hypertension [n (%)] | 11 (4.5%) |
| **SHS Exposure** |  |
| Ever Cabin SHS exposure [n (%)] | 139 (56.7%) |
| Cabin SHS exposure among exposed (years) | 16.6±9.1 |
| Time since last cabin SHS exposure (years) | 17.3±6.9 |
| Any form of non-cabin SHS exposure [n/N (%)] | 245/245 (100%) |
| Childhood home SHS exposure [n/N (%)] | 116/245 (47.3%) |
| Adult home SHS exposure [n/N (%)] | 59/245 (24.1%) |
| Non-airline occupational SHS exposure [n/N (%)] | 72/245 (29.4%) |
| Other SHS Exposure [n/N (%)] | 111/245 (45.3%) |
| **Symptoms** |  |
| mMRC Dyspnea Scale ≥1 [n (%)] | 25 (10.2%) |
| Participants with any respiratory symptoms [n (%)] | 112 (45.7%) |
| Participants ever experiencing shortness of breath [n (%)] | 29 (11.8%) |
| Participants with cough $\geq 2$ episodes/year [n (%)] | 103 (42%) |
| Participants with less activity than peers [n (%)] | 5 (2%) |
| **Pulmonary Function Tests** |  |
| FEV<sub>1</sub> (% predicted) | 103±12 |
| FVC (% predicted) | 107±13 |
| FEV<sub>1</sub>/FVC | 77±5 |
| FEV<sub>1</sub>/FVC (% predicted) | 95±6 |
| FEF<sub>25-75</sub> (% predicted) | 94±25 |
| FEF<sub>75</sub> (% predicted) | 112±44 |
| D<sub>LCO</sub> adjusted for Hgb (mL/min/mmHg) | 22.79±5.01 |
| D<sub>LCO</sub> adjusted for Hgb (% predicted) | 81±10 |
| D<sub>L</sub>/V<sub>A</sub> adjusted for Hgb (mL/min/mmHg/L) | 4.48±0.67 |
| D<sub>L</sub>/V<sub>A</sub> adjusted for Hgb (% predicted) | 123±16 |
| Alveolar volume (V<sub>A</sub>) (L) | 5.11±0.95 |
| Alveolar volume (V<sub>A</sub>) (L) (% predicted) | 95±12 |
| TLC (% predicted) | 103±11 |
| RV (% predicted) | 96±18 |
| RV/TLC | 33±7 |
| RV/TLC (% predicted) | 90±13 |
| FRC (% predicted) | 103±20 |
| FRC/TLC | 53±7 |
| FRC/TLC (% predicted) | 99±13 |

All participants had preserved spirometry (normal FEV_1_/FVC and FEV_1_ by LLN criterion) but some had mildly reduced diffusing capacity at 81±10 (median [IQR] <TOTAL range> of 80 [73-88]) <55-114> %predicted). The participants had FRC/TLC of 0.53±0.07 (at 99±13 %predicted; median [IQR] <TOTAL range> of 99 [91-106] <54-129> %predicted) and RV/TLC of 0.33±0.07 (at 90±13 %predicted; median [IQR] <TOTAL range> of 90 [82-98] <53-129> %predicted).

### Overall response to exercise

All participants reported ceasing exercise due to dyspnea or leg fatigue. Borg rating of perceived exercise scales were 4.2±1.9 for dyspnea, 5.1±1.9 for effort, and 4.7±1.9 for fatigue at peak exercise. When pressed to identify a single cause for ceasing exercise, 117 (48%) identified dyspnea as opposed to fatigue or effort as the main cause (Dyspnea_Peak_) (Table 2).

**Table 2.** Cardiopulmonary testing measurements. <u>Footnote</u>: Cardio-pulmonary measurements in participants who underwent exercise testing. Data are presented as mean ± standard deviation. Data are presented as mean ± standard deviation. Reference equations: percent predicted of normal values of cardiopulmonary outputs were calculated using Wassermann predicted formulas.^20^ Abbreviations-VO_2Peak_: peak oxygen uptake; VO_2Max.kg_: peak oxygen uptake per kilogram of body weight; Watts_Peak_: peak work stage completed in watts; VCO_2Peak_: peak carbon dioxide production; O_2_-Pulse_Peak_: oxygen uptake per heartbeat at peak exercise; VE_Peak_: peak minute ventilation value; RER: respiratory exchange ratio (VCO_2_/VO_2_) at peak exercise; RR_Peak_: peak respiratory rate; VT_Peak_: peak tidal volume; HR_Peak_: peak heart rate; HRR: heart rate reserve.

| **Cardiopulmonary exercise testing outputs** | **N=245** |
| --- | --- |
| Cardiopulmonary Testing Measurements |  |
| VO<sub>2Peak</sub> (L/min) | 1.557±0.476 |
| VO<sub>2Peak</sub> (% predicted) | 100±24 |
| Reached 100% predicted VO<sub>2Peak</sub> [n (%)] | 112 (45.7%) |
| VO<sub>2Peak</sub> <100% predicted | 131 (53.5%) |
| VO<sub>2Peak</sub> <84% predicted | 71 (29%) |
| VO<sub>2Peak</sub>/kg (mL/min.kg) | 23.82±7.02 |
| VO<sub>2Peak</sub>/kg (% predicted) | 85±18 |
| VO<sub>2Peak</sub>/kg < LLN [n (%)] | 33 (13.5%) |
| VCO<sub>2Peak</sub> (L/min) | 1.90±0.59 |
| Watts<sub>Peak</sub> (watts) | 156±46 |
| Watts<sub>Peak</sub> (% predicted) | 135±32 |
| Watts per stage (watts) | 27±8 |
| Stages completed | 6.0±1.3 |
| Symptoms at peak exercise (Borg Scale 0 to 10) |  |
| Shortness of Breath | 4.2±1.9 |
| Effort | 5.1±1.9 |
| Fatigue | 4.7±1.9 |
| Dyspnea<sub>Peak</sub> [n (%)] | 117 (47.8%) |
| Pulmonary Response |  |
| V<sub>EPeak</sub> (L/min) | 57.04±16.97 |
| V<sub>EPeak</sub> (% predicted) | 50±12 |
| RER | 1.21±0.13 |
| RR<sub>Peak</sub> (breaths/min) | 34.7±9.0 |
| $RR_{Peak} \geq 60$ [n/N (%)] | 1/235 (0.4%) |
| $V_{TPeak}$ (L) | $1.73 \pm 0.47$ |
| $V_{TPeak}$ (% predicted) | $86 \pm 17$ |
| $V_E/VO_{2Peak}$ | $37.1 \pm 6.2$ |
| $V_E/VCO_{2Peak}$ | $30.7 \pm 3.8$ |
| $V_E/VCO_{2Peak} \geq 60$ [n (%)] | 51 (20.8%) |
| $V_E/VCO_{2Peak}$ (% predicted) | $108 \pm 12$ |
| $V_E/VCO_{2Lowest}$ | $29.4 \pm 3.4$ |
| $V_E/VCO_{2Lowest}$ (% predicted) | $103 \pm 11$ |
| $VO_2$ at Anaerobic Threshold ($VO_{2AT}$) (L/min) | $0.99 \pm 0.33$ |
| $VO_2$ at Anaerobic Threshold ($VO_{2AT}$) (% $VO_{2Peak}$) | $63 \pm 13$ |
| $VO_{2AT} \leq 40\% VO_{2Peak}$ [n/N (%)] | 11/202 (5.4%) |
| $V_E/VCO_2$ at Anaerobic Threshold | $31.4 \pm 3.9$ |
| $V_E/VCO_2$ at Anaerobic Threshold (% predicted) | $103 \pm 11$ |
| Cardiovascular Response |  |
| $HR_{Baseline}$ (beat/min) | $67.5 \pm 12.4$ |
| $HR_{Peak}$ (beat/min) | $141.8 \pm 17.9$ |
| $HR_{Peak}$ (% predicted) | $85 \pm 10$ |
| $HR_{Peak} \leq 90\%$ predicted [n (%)] | 165 (67.3%) |
| HRR | $24 \pm 17$ |
| $HRR \geq 15$ [n (%)] | 176 (71.8%) |
| $HR_{Slope}$ (beat/min/10 Watts) | $4.73 \pm 1.19$ |
| $SBP_{Baseline}$ (mmHg) | $121.1 \pm 15.5$ |
| $SBP_{Peak}$ (mmHg) | $178.8 \pm 20.3$ |
| $SBP_{Slope}$ (mmHg/10 Watts) | $3.90 \pm 1.42$ |
| $DBP_{Baseline}$ (mmHg) | $76.5 \pm 8.9$ |
| DBP<sub>Peak</sub> (mmHg) | 93.1±9.8 |
| DBP<sub>Slope</sub> (mmHg/10 Watts) | 1.17±0.72 |
| HRE (met at least one criterion) [n/N (%)] | 149/226 (65.9%) |
| Rise in SBP ≥50 mmHg (≥60 mmHg for men) | 138/226 (61.1%) |
| SBP<sub>Peak</sub> ≥190 mmHg (≥210 mmHg for men) | 45/226 (19.9%) |
| DBP<sub>Peak</sub> ≥105 mmHg | 24/226 (10.6%) |
| O<sub>2</sub>-Pulse<sub>Peak</sub> (mL/beat) | 11.0±3.0 |
| O<sub>2</sub>-Pulse<sub>Peak</sub> (% predicted) | 116±25 |
| Reached 100% predicted O<sub>2</sub>-Pulse<sub>Peak</sub> [n (%)] | 181 (73.9%) |
| O<sub>2</sub>-Pulse<sub>Peak</sub><100% predicted [n (%)] | 64 (26.1%) |
| O<sub>2</sub>-Pulse<sub>Peak</sub>≤80% predicted [n (%)] | 8 (3.3%) |
| O<sub>2</sub>-Pulse<sub>Baseline</sub> (mL/beat) | 3.8±1.0 |
| O<sub>2</sub>-Pulse<sub>Slope</sub> (mL/beat/Watt) | 0.47±0.11 |
| O<sub>2</sub>-Pulse at Anaerobic Threshold (mL/beat) | 9.1±3.6 |
| O<sub>2</sub>-Pulse at Anaerobic Threshold (% O<sub>2</sub>-Pulse<sub>Peak</sub>) | 84±30 |
| SpO<sub>2Baseline</sub> | 99±1 |
| SpO<sub>2Peak</sub> | 98±1 |
| Change in SpO<sub>2</sub> at peak exercise | -0.6±1.4 |

### Pulmonary response to exercise

The average volume of oxygen consumption at peak exercise (VO_2Peak_) was 1,557±476 mL/min at 100±24 %predicted (median [IQR] <TOTAL range> VO_2Peak_ of 98 [82-113] <52-208> %predicted) with 71 (29%) having a VO_2Peak_ <84 %predicted, a presumed threshold for abnormal results. The volume of oxygen consumption by weight at peak exercise (VO_2Peak/kg_) was 23.8±7.0 mL.kg^-1^.min^-1^ at 85±18 %predicted (median [IQR] <TOTAL range> VO_2Peak/kg_ of 85 [72-94] <44-163> %predicted by FRIENDS reference equation)^28^ with 33 (14%) having a value below the LLN. This VO_2Peak_ was achieved at a peak workload of 156±46 watts (135±33 %predicted) (median [IQR] <TOTAL range> Watts_Peak_ of 135 [112-155] <64-273> %predicted). The ratio of oxy-hemoglobin to total hemoglobin (oxygen saturation or SpO_2Peak_) at peak exercise was 98±1% (median [IQR] <TOTAL range> SpO_2Peak_ of 99 [98-100] <94-100> %) with nearly all participants but one having a SpO_2Peak_ of ≥95% (one had SpO_2Peak_ of 94%, eleven had SpO_2Peak_ of 95%, and all other >95%).

The pulmonary response to exercise was also remarkable for a peak-exercise minute ventilation (V_EPeak_) of 57.0±17.0 L/min at 50±12 %predicted (median [IQR] <TOTAL range> of 50 [42-58] <18-90> %predicted) with only 12 (5%) exceeding the 70% threshold for inappropriate ventilatory response to maximum effort exercise. This level of minute ventilation at peak exercise was achieved through a tidal volumes at peak exercise (V_TPeak_) of 1.73±0.47 L at 86±17 %predicted (median [IQR] <TOTAL range> of 85 [76-96] <42-146> %predicted), and a peak-exercise respiratory rate (RR_Peak_) that remained below the 50 breaths/minute in 93% (218 out of 235) and below the 60 breaths/minute threshold in all but 1 participant (35±9 breaths/min; median [IQR] <TOTAL range> of 33 [28-40] <15-69> breaths/min) (Table 2). All participants reached their anaerobic threshold (VO_2AT_) as determined by V slope method. The VO_2AT_ was 994±327 mL/min at 63±13% of VO_2Peak_ (median [IQR] <TOTAL range> of 63 [56-71] <27-100> % of VO_2Peak_) (Table 2). The volume of oxygen consumption by weight at anaerobic threshold (VO_2AT/kg_) was17.8±4.9 mL.kg^-1^.min^-1^, which was 54±14 % of the maximum VO_2_ predicted (median [IQR] <TOTAL range> of 54 [43-63] <18-96>) % of the maximum VO_2_ predicted.

### Cardiovascular response to exercise

None of the participants reported any chest pain, chest tightness, lightheadedness, or dizziness during the CPET. None had any clinically significant electrocardiographic (ECG) changes or arrhythmia besides occasional premature ventricular contractions that did not increase in frequency with exercise testing. Nevertheless, many had abnormal cardiovascular response to exercise as described below.

Although only 11 (4%) of participants had reported history of hypertension (with their hypertension well-controlled), 66% percent of the participants (149 out of 226) showed hypertensive response to exercise by at least one established criterion (Table 2). The heart rate at peak exercise was 142±18 beat/min (85±10 %predicted; median [IQR] <TOTAL range> of 85 [79-93] <58-119> %predicted). Oxygen-pulse at peak exercise (O_2_-Pulse_Peak_) was 11.0±3.0 mL/beat (116±25 %predicted; median [IQR] <TOTAL range> of 113 [99-133] <64-224> %predicted), with 8 (3.3%) participants not achieving their 80% predicted values. (Table 2).

Furthermore, ventilatory efficiency (V_E_/VCO_2_) at peak exercise was 30.7±3.8 (108±12 %predicted; median [IQR] <TOTAL range> of 107 [99-114] <79-181> %predicted). The lowest V_E_/VCO_2_ was 29.4±3.4 (103±11 %predicted; median [IQR] <TOTAL range> of 104 [97-110] <45-129> %predicted) (Table 2).

### Association of exercise capacity with cardiovascular response measures, air trapping, and SHS exposure

In models that included each (single) cardiopulmonary exercise testing (CPET) output separately as a predictor along with age, sex, height, and BMI as covariates, workload (as measured by Watt_Peak_ or VO_2Peak_) and cumulative work (as measured by Watt-Minute) were directly associated with the peak of heart rate and systolic blood pressure during exercise (HR_Peak_ and SBP_Peak_). However, workload and cumulative work were inversely associated with the rate of increase in SBP, DBP, and HR (HR_Slope_, SBP_Slope_, and DBP_Slope_) (all P<0.05), with faster rise of these indices being associated with lower workload or cumulative work the participants could achieve (Table 3). Similarly, workload (Watt_Peak_) and cumulative work (Watt-Minute) were associated directly with O_2_-Pulse_Peak_ but inversely with O_2_-Pulse_Slope_. Moreover, Watt_Peak_, VO_2Peak_, and Watt-Minute were directly associated with respiratory rate, tidal volume, and minute ventilation at peak exercise (RR_Peak_, V_TPeak_, V_EPeak_) and inversely with a lung function measure of air trapping (RV/TLC) (all P<0.05) (Table 3).

**Table 3.** Associations of exercise capacity with each of the cardiopulmonary exercise outputs or SHS exposure. <u>Footnote</u>: The association between exercise capacity (dependent variable) and each of the cardiopulmonary exercise outputs, pulmonary outputs, or SHS exposure (independent variable) were individually assessed by linear regression modeling with adjustment for age, sex, height, BMI, and corresponding baseline values (for slope variables). Statistical significance was determined by a P value<0.05. * The associations of VO2Peak with O2-PulsePeak and O2-PulseSlope were not computed due to existence of “mathematical coupling” between VO_2_ and O_2_-Pulse, which depends on VO_2_ for its calculation (O_2_-Pulse=VO_2_/HR). Abbreviations-VO_2Peak_: peak oxygen uptake; SHS: secondhand smoke, O_2_-Pulse_Peak_: oxygen uptake per heartbeat at peak exercise; HR: heart rate; SBP: systolic blood pressure; DBP: diastolic blood pressure; RR_Peak_: peak respiratory rate; VE_Peak_: peak minute ventilation value; VT_Peak_: peak tidal volume; VO_2AT_: oxygen uptake at anaerobic threshold; VCO_2Peak_: peak carbon dioxide production; RV: residual volume; TLC: total lung capacity; FRC: functional residual capacity.

|  | Dependent variables |  |  |  |  |  |  |  |  |
| --- | --- | --- | --- | --- | --- | --- | --- | --- | --- |
|  | Peak Workload (watt)<br>(Watts<sub>Peak</sub>) |  |  | Cumulative Work (watt-minute)<br>(Work<sub>Total</sub>) |  |  | Peak volume of O<sub>2</sub> consumption<br>(mL/min) (VO<sub>2Peak</sub>) |  |  |
| Independent variables | N | PE±SEM<br>95% CI | P value | N | OR±SEM<br>95% CI | P value | N | OR±SEM<br>95% CI | P value |
| **Cardiovascular outputs** |  |  |  |  |  |  |  |  |  |
| **Peaks** |  |  |  |  |  |  |  |  |  |
| O<sub>2</sub>-Pulse<sub>Peak</sub> | 245 | 10.3±0.7<br>(8.8 to 11.7) | <0.001 | 239 | 53.9±4.6<br>(44.9 to 62.9) | <0.001 | N/A * |  |  |
| HR<sub>Peak</sub> | 245 | 0.7±0.1<br>(0.4 to 0.9) | <0.001 | 239 | 4.4±0.7<br>(3.1 to 5.6) | <0.001 | 245 | 8.0±1.1<br>(5.7 to 10.2) | <0.001 |
| SBP<sub>Peak</sub> | 226 | 0.3±0.1<br>(0.1 to 0.5) | 0.004 | 224 | 2.2±0.6<br>(1.0 to 3.5) | <0.001 | 226 | 3.1±1.1<br>(0.9 to 5.4) | 0.006 |
| DBP<sub>Peak</sub> | 226 | 0.1±0.2<br>(-0.3 to 0.5) | 0.621 | 224 | 0.1±1.2<br>(-2.4 to 2.5) | 0.945 | 226 | 0.2±2.2<br>(-4.2 to 4.6) | 0.923 |
| **Slopes** |  |  |  |  |  |  |  |  |  |
| O<sub>2</sub>-Pulse<sub>Slope</sub> | 238 | -64.5±18.1<br>(-100.1 to -28.9) | <0.001 | 238 | -251.9±109.5<br>(-467.6 to -36.2) | 0.022 | N/A * |  |  |
| HR<sub>Slope</sub> | 238 | -11.7±1.9<br>(-15.5 to -8.0) | <0.001 | 238 | -40.9±11.7<br>(-63.9 to -17.8) | <0.001 | 238 | -64.1±20.9<br>(-105.2 to -23.0) | 0.002 |
| SBP<sub>Slope</sub> | 227 | -7.0±1.5<br>(-10.1 to -4.0) | <0.001 | 225 | -22.3±9.3<br>(-40.7 to -4.0) | 0.017 | 227 | -44.1±16.6<br>(-76.8 to -11.4) | 0.008 |
| DBP<sub>Slope</sub> | 227 | -12.7±3.0<br>(-18.6 to -6.9) | <0.001 | 225 | -50.0±17.7<br>(-84.9 to -15.1) | 0.005 | 227 | -113.7±31.3<br>(-175.5 to -52.0) | <0.001 |
| **Pulmonary outputs** |  |  |  |  |  |  |  |  |  |
| RR<sub>Peak</sub> | 235 | 1.1±0.2<br>(0.6 to 1.5) | <0.001 | 233 | 7.9±1.3<br>(5.4 to 10.4) | <0.001 | 235 | 17.9±2.1<br>(13.8 to 22.1) | <0.001 |
| VE<sub>Peak</sub> | 245 | 1.4±0.1<br>(1.2 to 1.7) | <0.001 | 239 | 8.4±0.6<br>(7.2 to 9.6) | <0.001 | 245 | 17.5±0.9<br>(15.7 to 19.3) | <0.001 |
| VT<sub>Peak</sub> | 239 | 37.2±5.6<br>(26.1 to 48.3) | <0.001 | 237 | 177.6±33.5<br>(111.5 to 243.6) | <0.001 | 239 | 307.8±59.2<br>(191.1 to 424.5) | <0.001 |
| VO<sub>2AT</sub> | 202 | 57.8±7.7<br>(42.6 to 73.0) | <0.001 | 200 | 287.1±47.4<br>(193.6 to 380.5) | <0.001 | 202 | 799.3±71.3<br>(658.8 to 939.9) | <0.001 |
| **VE/VCO<sub>2Peak</sub>** | **245** | **-1.2±0.6**<br>**(-2.3 to -0.02)** | **0.046** | 239 | -0.4±3.4<br>(-7.2 to 6.4) | 0.906 | 245 | -7.9±6.1<br>(-19.9 to 4.0) | 0.191 |
| **RV/TLC** | **235** | **-1.6±0.5**<br>**(-2.5 to -0.6)** | **0.001** | **230** | **-8.6±2.8**<br>**(-14.1 to -3.1)** | **0.002** | **235** | **-14.3±5.0**<br>**(-24.2 to -4.4)** | **0.004** |
| **FRC/TLC** | 220 | -0.3±0.4<br>(-1.1 to 0.4) | 0.363 | 219 | -2.1±2.2<br>(-6.5 to 2.2) | 0.337 | 220 | -5.0±4.0<br>(-12.8 to 2.8) | 0.210 |
| **SHS Exposure** |  |  |  |  |  |  |  |  |  |
| **Airline SHS exposure (years)** | **241** | **-0.8±0.2**<br>**(-1.2 to -0.3)** | **<0.001** | 235 | -2.3±1.4<br>(-5.1 to 0.4) | 0.092 | **241** | **-5.0±2.4**<br>**(-9.7 to -0.2)** | **0.040** |
| **Childhood home SHS exposure (Y/N)** | 245 | -0.5±4.1<br>(-8.7 to 7.6) | 0.895 | 239 | 4.7±24.3<br>(-43.1 to 52.5) | 0.845 | 245 | -17.7±42.5<br>(-101.4 to 65.9) | 0.676 |
| **Adult home SHS exposure (Y/N)** | 245 | -5.1±4.9<br>(-14.7 to 4.5) | 0.292 | 239 | -19.0±28.8<br>(-75.7 to 37.6) | 0.509 | 245 | -50.9±50.2<br>(-149.7 to 47.9) | 0.311 |

Workload (Watt_Peak_ and VO_2Peak_) was also inversely associated with years of exposure to cabin SHS (both P<0.05), but was not significantly associated with non-cabin SHS exposure including childhood and adulthood home (Table 3).

### Association of respiratory symptoms with cardiopulmonary response measures

As shown in Table 4, baseline respiratory symptoms (as measured by mMRC) were inversely associated with workload (Watt_Peak_ or VO_2Peak_) and cumulative work (Watt-Minute) achieved. Baseline respiratory symptoms at rest (as measured by FAMRI questionnaire) and at peak exercise (as measured by Borg Scale) were not significantly associated with exercise capacity.

**Table 4.**
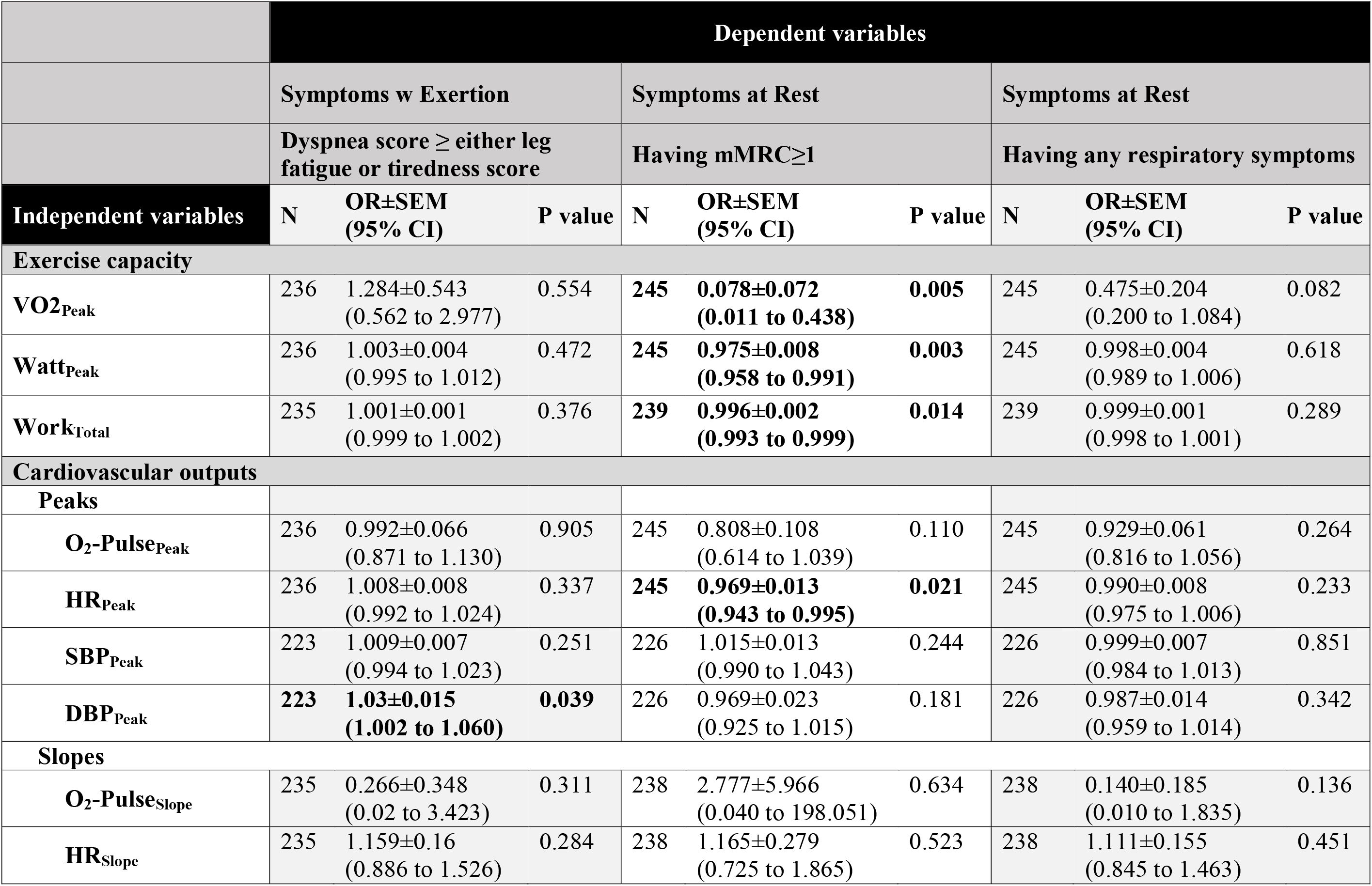
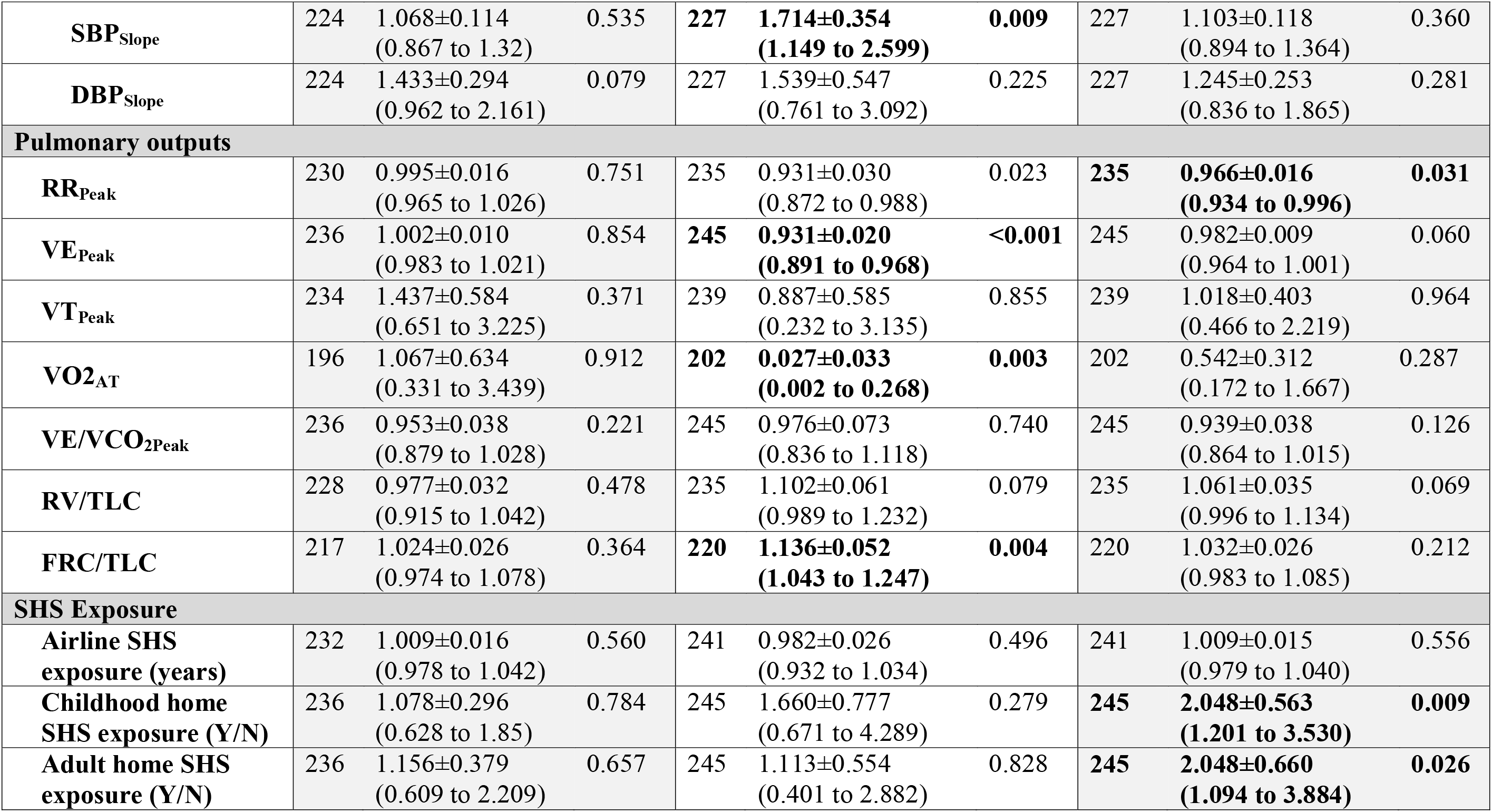
Associations of respiratory symptoms with each of the cardiopulmonary exercise outputs and SHS exposure. <u>Footnote</u>: Associations of respiratory symptoms at rest (mMRC and FAMRI questionnaires) and during exercise (Dyspnea score) (dependent variables) with each of the cardiopulmonary exercise outputs and SHS exposure (predictors) were assessed by univariate linear regression models with adjustment for age, sex, height, BMI, and corresponding baseline values (for slope predictors). Statistical significance was determined by a P value<0.05. Abbreviations-VO_2Peak_: peak oxygen uptake; SHS: secondhand smoke, O_2_-Pulse_Peak_: oxygen uptake per heartbeat at peak exercise; HR: heart rate; SBP: systolic blood pressure; DBP: diastolic blood pressure; RR_Peak_: peak respiratory rate; VE_Peak_: peak minute ventilation value; VT_Peak_: peak tidal volume; VO_2AT_: oxygen uptake at Anaerobic Threshold; VCO_2Peak_: peak carbon dioxide production; RV: residual volume; TLC: total lung capacity; FRC: functional residual capacity.

Overall, only a few CPET outputs were associated with respiratory symptoms. Among cardiovascular outputs, the likelihood of having an mMRC≥1 was associated with higher rise in SBP (SBP_Slope_) and with lower HR at peak exercise (HR_Peak_). On the other hand, the likelihood of stopping exercise due to dyspnea as measured by BORG scale was associated with a higher DBP at peak exercise (DBP_Peak_). Among pulmonary outputs, the likelihood of having an mMRC≥1 was associated with lower RR and V_E_ at peak exercise (RR_Peak_ and V_EPeak_), and lower anaerobic threshold (VO_2AT_). Furthermore, baseline respiratory symptoms were significantly associated with FRC/TLC with more air trapping being associated with higher likelihood of having respiratory symptoms at baseline (RV/TLC association was marginally significant; P≤0.079).

### Association of cardiovascular response measures with SHS exposure

Among the cardiovascular predictors of exercise capacity, O_2_-Pulse_Peak_ was inversely associated with years of exposure to cabin SHS, showing a parameter estimate of −0.032±0.015 (P=0.040), which indicates a decrease in O_2_-Pulse_Peak_ of 0.32 mL/beat for every 10 years increased in exposure to cabin SHS (Table 7). Consistently, the rate of increase in heart rate (HR_Slope_) was directly associated with years of exposure to cabin SHS. Furthermore, Mediation analysis showed that a substantial fraction of cabin SHS exposure association with exercise capacity (Watt_Peak_) was mediated through SHS effect on O_2_-Pulse (41%; P=0.038) (Table 6). Although not significant, cabin SHS association with cumulative work (Watt-Minute) also seemed to be mediated through O_2_-Pulse (74%; P=0.112).

**Table 5.** Mediation analysis of effect of SHS exposure on exercise capacity through mediators. <u>Footnote</u>: To see if the association of SHS exposure (independent variable) with exercise capacity (dependent variable) was mediated through the cardiovascular or pulmonary outputs, mediation analyses were performed with adjustment for age, sex, height, and BMI and inclusion of one mediator at a time. Statistical significance was determined by a P value<0.05. * The associations of VO_2Peak_ with O2-Pulse_Peak_ and O2-Pulse_Slope_ were not computed due to existence of “mathematical coupling” between VO_2_ and O_2_-Pulse, which depends on VO_2_ for its calculation (O_2_-Pulse=VO_2_/HR). Abbreviations-VO_2Peak_: peak oxygen uptake; SHS: secondhand smoke; O_2_-Pulse_Peak_: oxygen uptake per heartbeat at peak exercise; HR: heart rate; SBP: systolic blood pressure; DBP: diastolic blood pressure; RR_Peak_: peak respiratory rate; VE_Peak_: peak minute ventilation value; VT_Peak_: peak tidal volume; RV: residual volume; TLC: total lung capacity; FRC: functional residual capacity.

|  | Mediation analysis of effect of SHS exposure to |  |  |  |  |  |  |  |  |
| --- | --- | --- | --- | --- | --- | --- | --- | --- | --- |
|  | Watts<sub>Peak</sub> |  |  | Work<sub>Total</sub> |  |  | VO<sub>2Peak</sub> |  |  |
| Mediators | N | % mediated | P value | N | % mediated | P value | N | % mediated | P value |
| **Cardiovascular outputs** |  |  |  |  |  |  |  |  |  |
| **Peaks** |  |  |  |  |  |  |  |  |  |
| **O<sub>2</sub>-Pulse<sub>Peak</sub>** | 241 | 40.8(4.4 to 76.2) | 0.038 | 235 | 73.6(-170.7 to 319.8) | 0.112 |  | N/A * |  |
| **HR<sub>Peak</sub>** | 241 | 0.03(-33.5 to 21.5) | 0.998 | 235 | 8.0(-106.2 to 112.8) | 0.750 | 241 | 1.6(-109.5 to 55.6) | 0.962 |
| **SBP<sub>Peak</sub>** | 223 | 2.5(-17.4 to 23) | 0.718 | 221 | 4.8(-74.5 to 79.9) | 0.736 | 223 | 3.2(-53.4 to 42.3) | 0.726 |
| **DBP<sub>Peak</sub>** | 223 | 0.2(-11.9 to 12.9) | 0.892 | 221 | 0.5(-28.8 to 47.5) | 0.884 | 223 | 0.3(-20.5 to 32) | 0.894 |
| **Slopes** |  |  |  |  |  |  |  |  |  |
| **O<sub>2</sub>-Pulse<sub>Slope</sub>** | 234 | 11.7(-7.4 to 37.9) | 0.214 | 234 | 14.1(-81.6 to 101.9) | 0.278 |  | N/A * |  |
| **HR<sub>Slope</sub>** | 234 | 17.6(-0.6 to 49.7) | 0.058 | 234 | 18.9(-80.6 to 138.7) | 0.160 | 234 | 13.2(-4.6 to 75.4) | 0.114 |
| **SBP<sub>Slope</sub>** | 224 | 1.7(-30.5 to 24) | 0.848 | 222 | 0.7(-53.9 to 38.4) | 0.868 | 224 | 1.0(-53.8 to 30.8) | 0.882 |
| **DBP<sub>Slope</sub>** | 224 | 1.9(-31.1 to 25) | 0.840 | 222 | 0.7(-83.2 to 44.9) | 0.934 | 224 | 2.1(-75.4 to 39.1) | 0.852 |
| **Pulmonary outputs** |  |  |  |  |  |  |  |  |  |
| **RR<sub>Peak</sub>** | 231 | 7.3(-19.9 to 34) | 0.474 | 229 | 26.3(-167.8 to 217.8) | 0.294 | 231 | 18.1(-82.2 to 90.2) | 0.462 |
| **VE<sub>Peak</sub>** | 241 | 30.0(-13.4 to 61.7) | 0.116 | 235 | 70.4(-166.1 to 277.7) | 0.116 | 241 | 58.9(-47.9 to 156.9) | 0.106 |
| **VT<sub>Peak</sub>** | 235 | 11.5(-17.4 to 38.8) | 0.300 | 233 | 14.8(-61.5 to 124.4) | 0.398 | 235 | 13.7(-35.4 to 74.9) | 0.316 |
| **RV/TLC** | 234 | 7.3(-7 to 42.2) | 0.284 | 229 | 10.5(-59.2 to 134.9) | 0.310 | 234 | 8.1(-14.3 to 99.9) | 0.318 |
| **FRC/TLC** | 219 | 4.3(-4.8 to 31.1) | 0.276 | 218 | 5.2(-30.1 to 73.1) | 0.366 | 219 | 6.6(-9.7 to 83.5) | 0.256 |

**Table 6.**
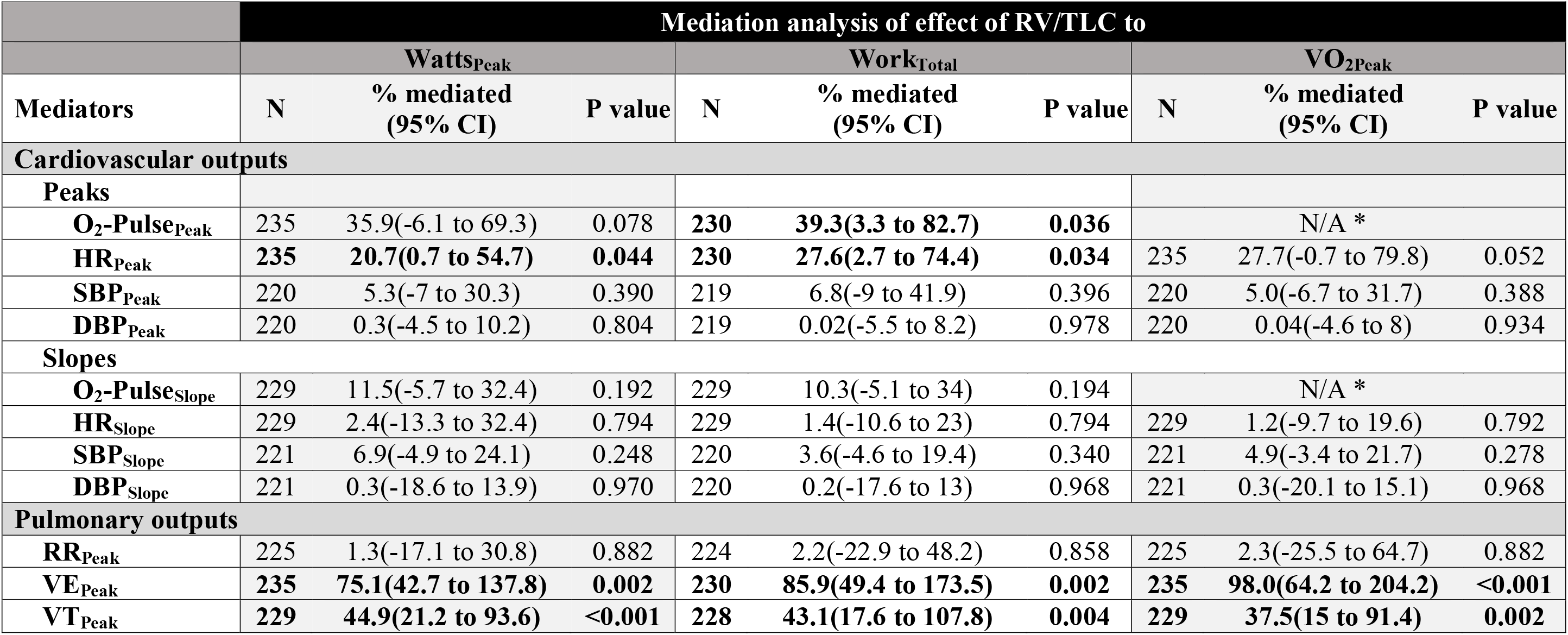
Mediation analysis of effect of air trapping on exercise capacity through mediators. <u>Footnote</u>: To see if the association of air trapping (independent variable) with exercise capacity (dependent variable) was mediated through the cardiovascular or pulmonary outputs, mediation analyses were performed with adjustment for age, sex, height, and BMI and inclusion of one mediator at a time. Statistical significance was determined by a P value<0.05. * The associations of VO_2Peak_ with O2-Pulse_Peak_ and O2-Pulse_Slope_ were not computed due to existence of “mathematical coupling” between VO_2_ and O_2_-Pulse, which depends on VO_2_ for its calculation (O_2_-Pulse=VO_2_/HR). Abbreviations-VO_2Peak_: peak oxygen uptake; SHS: secondhand smoke; O_2_-Pulse_Peak_: oxygen uptake per heartbeat at peak exercise; HR: heart rate; SBP: systolic blood pressure; DBP: diastolic blood pressure; RR_Peak_: peak respiratory rate; VE_Peak_: peak minute ventilation value; VT_Peak_: peak tidal volume; RV: residual volume; TLC: total lung capacity.

|  | Mediation analysis of effect of RV/TLC to |  |  |  |  |  |  |  |  |
| --- | --- | --- | --- | --- | --- | --- | --- | --- | --- |
|  | Watts<sub>Peak</sub> |  |  | Work<sub>Total</sub> |  |  | VO<sub>2Peak</sub> |  |  |
| Mediators | N | % mediated<br>(95% CI) | P value | N | % mediated<br>(95% CI) | P value | N | % mediated<br>(95% CI) | P value |
| **Cardiovascular outputs** |  |  |  |  |  |  |  |  |  |
| **Peaks** |  |  |  |  |  |  |  |  |  |
| **O<sub>2</sub>-Pulse<sub>Peak</sub>** | 235 | 35.9(-6.1 to 69.3) | 0.078 | **230** | **39.3(3.3 to 82.7)** | **0.036** |  | N/A * |  |
| **HR<sub>Peak</sub>** | **235** | **20.7(0.7 to 54.7)** | **0.044** | **230** | **27.6(2.7 to 74.4)** | **0.034** | 235 | 27.7(-0.7 to 79.8) | 0.052 |
| **SBP<sub>Peak</sub>** | 220 | 5.3(-7 to 30.3) | 0.390 | 219 | 6.8(-9 to 41.9) | 0.396 | 220 | 5.0(-6.7 to 31.7) | 0.388 |
| **DBP<sub>Peak</sub>** | 220 | 0.3(-4.5 to 10.2) | 0.804 | 219 | 0.02(-5.5 to 8.2) | 0.978 | 220 | 0.04(-4.6 to 8) | 0.934 |
| **Slopes** |  |  |  |  |  |  |  |  |  |
| **O<sub>2</sub>-Pulse<sub>Slope</sub>** | 229 | 11.5(-5.7 to 32.4) | 0.192 | 229 | 10.3(-5.1 to 34) | 0.194 |  | N/A * |  |
| **HR<sub>Slope</sub>** | 229 | 2.4(-13.3 to 32.4) | 0.794 | 229 | 1.4(-10.6 to 23) | 0.794 | 229 | 1.2(-9.7 to 19.6) | 0.792 |
| **SBP<sub>Slope</sub>** | 221 | 6.9(-4.9 to 24.1) | 0.248 | 220 | 3.6(-4.6 to 19.4) | 0.340 | 221 | 4.9(-3.4 to 21.7) | 0.278 |
| **DBP<sub>Slope</sub>** | 221 | 0.3(-18.6 to 13.9) | 0.970 | 220 | 0.2(-17.6 to 13) | 0.968 | 221 | 0.3(-20.1 to 15.1) | 0.968 |
| **Pulmonary outputs** |  |  |  |  |  |  |  |  |  |
| **RR<sub>Peak</sub>** | 225 | 1.3(-17.1 to 30.8) | 0.882 | 224 | 2.2(-22.9 to 48.2) | 0.858 | 225 | 2.3(-25.5 to 64.7) | 0.882 |
| **VE<sub>Peak</sub>** | **235** | **75.1(42.7 to 137.8)** | **0.002** | **230** | **85.9(49.4 to 173.5)** | **0.002** | **235** | **98.0(64.2 to 204.2)** | **<0.001** |
| **VT<sub>Peak</sub>** | **229** | **44.9(21.2 to 93.6)** | **<0.001** | **228** | **43.1(17.6 to 107.8)** | **0.004** | **229** | **37.5(15 to 91.4)** | **0.002** |

### Cardiovascular response measures and baseline air trapping

Given the physical location of lung, heart, and great vasculature within thoracic cavity, we examined the potential interaction of baseline air trapping and cardiovascular outputs of exercise. To do this, we examined whether the cardiovascular predictors of exercise capacity including O_2_-Pulse (proxy of stroke volume) and blood pressure (SBP and DBP) mediate the association of air trapping (FRC/TLC and RV/TLC) with exercise capacity.

These mediation analyses showed that a substantial fraction of RV/TLC association with exercise capacity (cumulative work [Watt-Minute]) was mediated through RV/TLC effect on O_2_-Pulse at peak exercise (39%; P=0.036) (Table 6). Although not significant, RV/TLC association with workload (Watt_Peak_) also seemed to be mediated through O_2_-Pulse (36%; P=0.078). Consistent with this finding, the heart rate at peak exercise (HR_Peak_) also mediated the association of RV/TLC with exercise capacity (mediated 21% and 28% of the total effect of RV/TLC on Watts_Peak_ and Work_Total_, respectively; all P<0.05).

## Discussion

In this observational study of a never-smoking cohort with a history of remote but prolonged exposure to SHS, we found the cohort to have an abnormal cardiovascular response to exercise that was proportional to their SHS exposure. Exercise capacity, as measured by highest workload completed (Watts_Peak_) or volume of oxygen uptake at peak exercise (VO_2Peak_), was associated with years of exposure to SHS. Remarkably, over 40% of the association of exercise capacity (Watts_Peak_) with SHS was dependent on O_2_-Pulse_Peak_, which suggests that the effect of SHS exposure on exercise capacity is mediated through SHS effect on stroke volume and cardiac output. We also found suggestive evidence, although not statistically significant (P=0.078), that pulmonary air trapping (elevated RV/TLC) contributes to lower exercise capacity through its effect on O_2_-Pulse_Peak_, implicating an interacting lung and heart pathophysiology between pulmonary hyperinflation and cardiac output that further impairs exercise capacity (Figure 2). Furthermore, we found over 60% of the participants to have a hypertensive response to exercise, suggesting that abnormal escalation of afterload contributed to lower exercise capacity in this SHS-exposed cohort in whom, with the exception of a few (4.3%) with well-controlled hypertension, none had any known hypertensive or cardiovascular disease.

**Figure 1.**
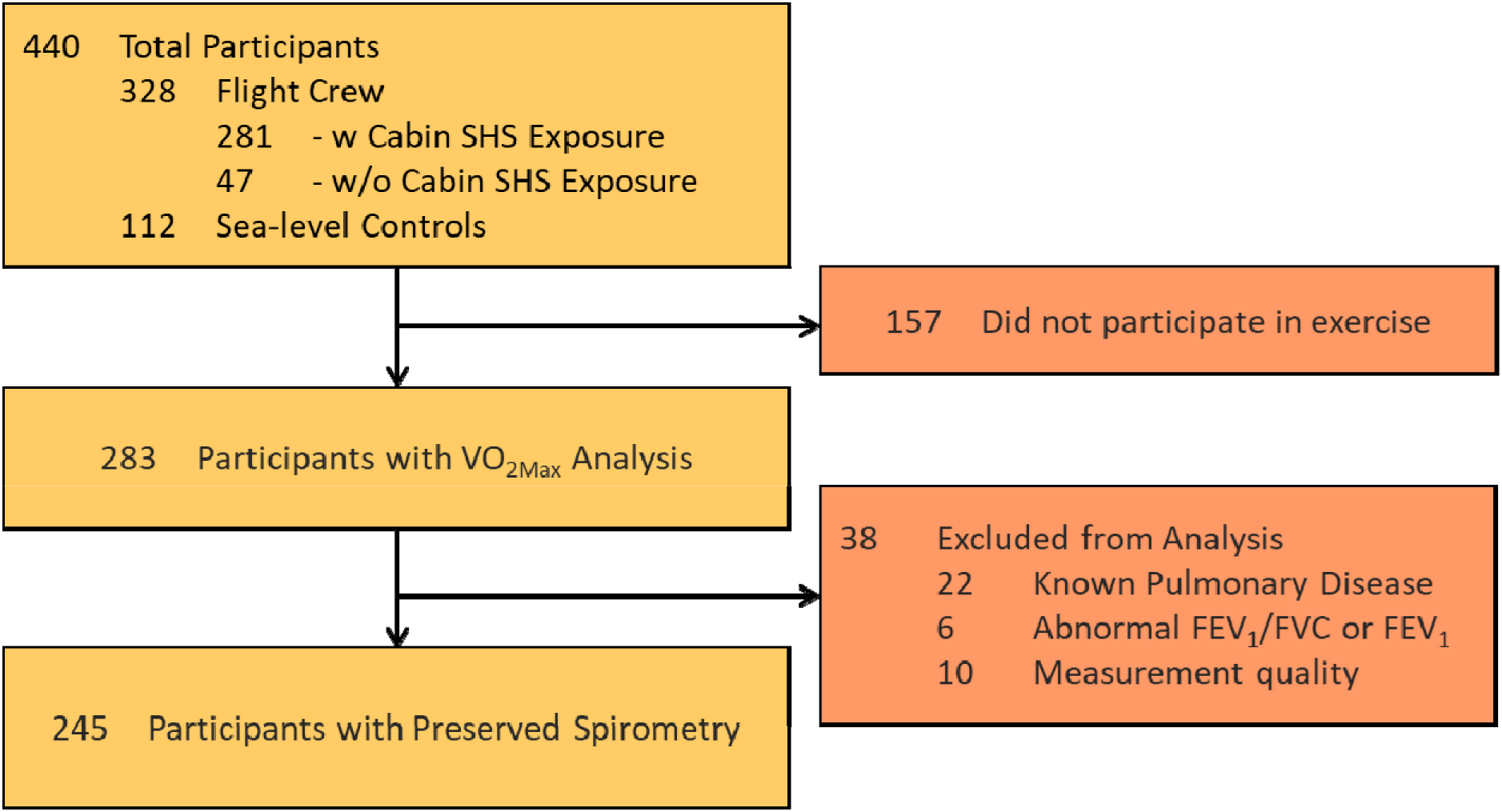
Participants flow through the study. Abbreviations-SHS=secondhand tobacco smoke; PFT= pulmonary function test; FEV1= forced expiratory volume in 1 second; FVC= forced vital capacity; BMI= body mass index; VO_2Max_= volume of maximum oxygen uptake.

**Figure 2.**
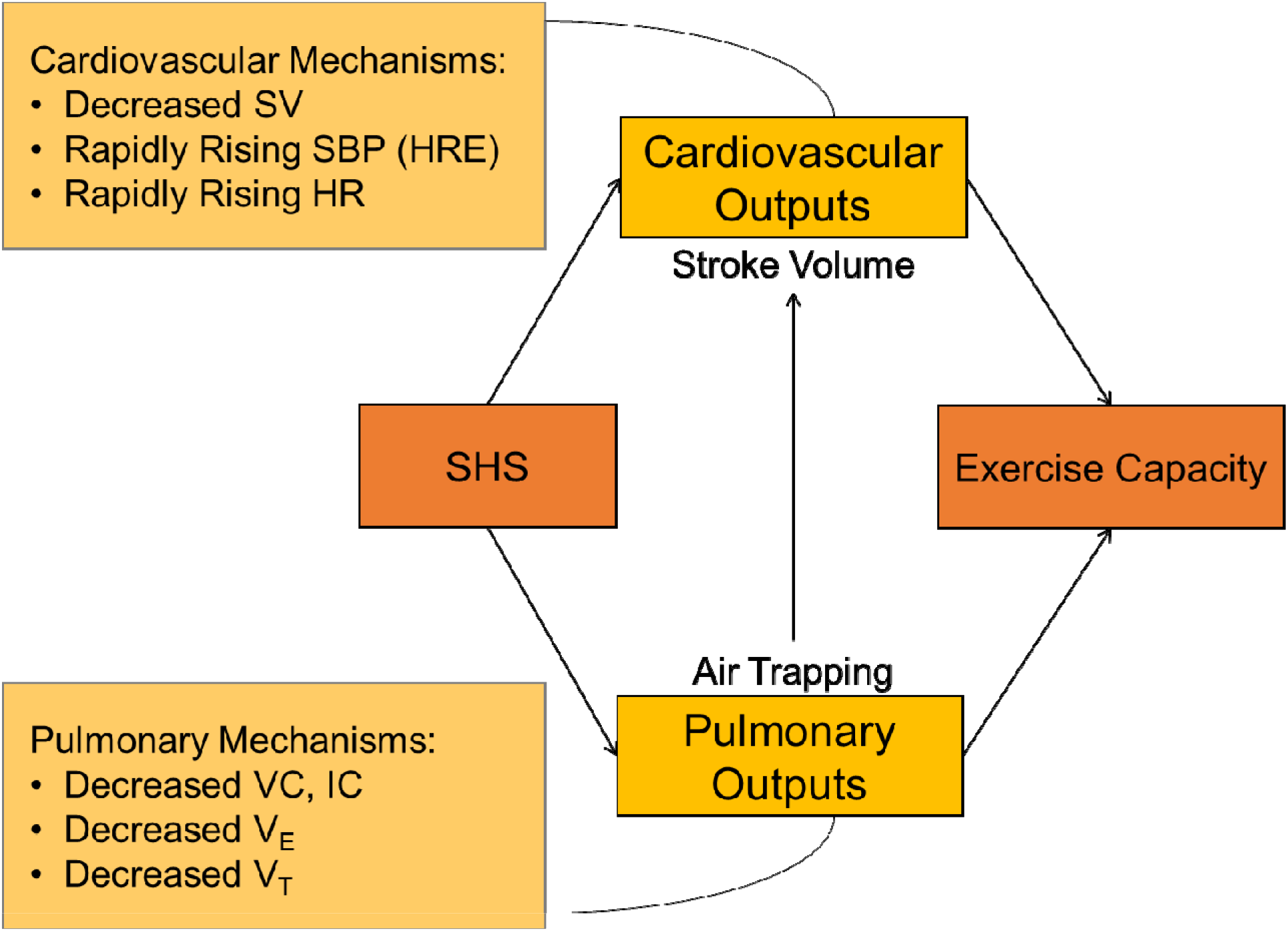
Proposed model referenced in the discussion. Illustration of mediation effects between SHS exposure and exercise capacity. Abbreviations-SV= cardiac stroke volume; HRE= hypertensive response to exercise; HR= heart rate; SHS=secondhand tobacco smoke; VC= vital capacity; IC= inspiratory capacity; V_E_= minute ventilation; V_T_= volume of tidal breathing.

Overall, we found evidence that prolonged exposure to SHS, even when remote, is associated with cardiovascular abnormalities suggestive of occult cardiovascular dysfunction with potential additional contribution from pulmonary hyperinflation. These abnormalities reveal subtle but lower cardiopulmonary functional reserve, manifested here as lower exercise capacity, and implicate a reduced efficiency of the body’s oxygen delivery machinery, which could be disadvantageous during the times of increased cardiopulmonary output demands as in physiological distress or disease.

In previous studies, we have shown this cohort of never-smokers with a history of prolonged remote SHS exposure to have abnormal lung function at rest and abnormal pulmonary response to exercise including (1) reduced diffusing capacity at rest,^13^ (2) reduced pulmonary capillary recruitment (as measured by impaired rise in diffusing capacity) during exercise,^14^ (3) decreased small airways airflow indices on spirometry (maximal flow in mid-and end-expiratory airflows [FEF_25-75%_ and FEF_75%_]),^13^ (4) plethysmographic and radiographic evidence of pulmonary air trapping at rest,^15^ and (5) progressive (dynamic) pulmonary hyperinflation during exercise.^15^ Overall, these abnormalities are suggestive of presence of an unrecognized early or mild obstructive lung disease that, while not meeting the spirometric definition of COPD, is consistent with an early/mild disease that could be categorized as “pre-COPD” and could contribute to lower pulmonary reserve and potential adverse health outcomes.^29, 30^

In the current study, we sought to determine whether there are also subtle cardiovascular abnormalities in this SHS-exposed cohort that are indicative of an occult and/or subclinical cardiovascular disease. Our finding that past exposure to SHS is a predictor of exercise capacity in an O_2_-pulse-dependent (a proxy of stroke volume and cardiac output) manner is novel and suggests that SHS exposure has lasting effect on cardiac function that is observable years after the exposure has ceased, and is consistent with recent findings that long-standing *direct* tobacco smoking impairs cardiac systolic function as evident by an increase in left and right ventricle end-systolic volumes and reduced left and right ejection fractions.^31^ Although it remains unclear how exposure to tobacco smoke, *direct* or *indirect*, causes an impairment in cardiac function, an interaction between pulmonary and cardiovascular systems, which occupy the same body cavity (thorax), has been proposed to play a role. Air trapping as measured by lung volumes (FRC/TLC and RV/TLC) is the earliest manifestation of COPD ^29, 30^ and is associated with reduced exercise capacity due to ventilatory limitation caused by progressive air trapping and pulmonary hyperinflation.^15^ Changes in lung volumes due to pulmonary hyperinflation could cause increased intra-thoracic pressures, particularly during exertion, and thus adversely affect the cardiovascular function.^23, 32^ To investigate this possible mechanism, we examined whether air trapping did contribute to exercise capacity through an interaction with cardiac output by performing a mediation analysis among pulmonary air trapping (FRC/TLC and RV/TLC), cardiac stroke volume (O_2_-pulse), and exercise capacity (VO_2Peak_) (Figure 2). Although the analysis did not reach statistical significance (P=0.078 for RV/TLC) and thus could not provide any further corroborating evidence for our hypothesis, the analysis did suggest that a substantial proportion (36%) of air trapping effect on VO_2Peak_ may be mediated through stroke volume. Further research in this area would be needed to better understand the interaction of cardiovascular and pulmonary systems in the context of exposure to direct or indirect smoke.

Another remarkable observation in our study was the common presence of hypertensive response to exercise (HRE). Despite some data to the contrary,^33, 34^ studies have described HRE in individuals with or without presence of baseline hypertension to be associated with impaired systolic and/or diastolic dysfunction even in the setting of having negative stress test and preserved left ventricular ejection fraction.^35, 36^ In fact, increasing evidence indicates that HRE is associated with functional and structural heart abnormalities, future development of hypertension, and increased cardiovascular morbidity and mortality.^37–39^ In our study, we found a substantial number of participants (62%) to have HRE despite all either having no history of hypertension (95%) or having hypertension that was well-controlled (5%). Studies have reported the prevalence of HRE to be between 3% to 40% among apparently healthy cohorts of varying age, sex, and ethnicity.^40–42^ Although we did not observe a significant association between presence of HRE and history of SHS exposure, the high prevalence of HRE among this never-smoking but SHS-exposed healthy cohort is remarkable, and suggests a potential pathogenetic role for SHS exposure in development of HRE.

### Limitations

Our study has limitations that should be kept in view. First, there may be concerns about the generalizability of the findings because the cohort studied are mostly women, which reflects the demographics of those who worked in airlines as flight crew in the latter half of the last century when smoking in aircraft cabin was permitted. The choice to study the flight crew allowed for overcoming the challenge of long-term SHS exposure assessment by providing the ability to generate a more objective and reproducible exposure index based on employment history and the smoking ban timeline on domestic and international flights of different airlines.^13^ Women have been reported to be more susceptible to adverse health effects of tobacco smoke.^43^ However, at the minimum, the findings should be generalizable to women. Second, the cardiovascular findings reported in this study are mainly derived from CPET with no imaging (such as echocardiography and magnetic resonance imaging [MRI]) or other clinical studies to provide additional robust evidence to corroborate our findings. Such studies are needed and are in progress (ClinicalTrials.gov Identifier: NCT04715568). Nevertheless, studies describing cardiovascular health effects of *direct* smoking using echocardiography and MRI have been previously reported, which corroborate our findings.^44, 45^ Our report however is the first report that describes the chronic and long-term cardiovascular health effects due to past prolonged exposure to SHS. Third, while we found association of exercise capacity (Watts_Peak_) with years of SHS exposure, the association of respiratory symptoms with SHS was less striking and less consistent across the different questionnaire platforms. However, it is not uncommon to see differences in scores across different respiratory questionnaires,^46^ and similarly, baseline respiratory symptoms (mMRC and UCSF FAMRI SHS questionnaires) may measure different things and thus produce different scores compared to those done at peak exercise (Borg Scale). For example, participants who had impairments at baseline and thus were more symptomatic are likely to not perform as well during the exercise and thus may report lesser symptoms in a sub-maximal effort exercise test.

## Conclusion

Healthy never-smokers with history of remote but prolonged exposure to SHS have an abnormal cardiovascular response to exercise, which is characterized by a stroke volume (oxygen-pulse) and thus an exercise capacity that are reduced proportional to their years of exposure to SHS. The mechanisms by which past exposure to SHS may limit stroke volume and thus exercise capacity is unknown, but pulmonary hyperinflation may have a contributory effect beyond its role in ventilatory limitation. Furthermore, these SHS-exposed individuals have a considerably higher prevalence of hypertensive response to exercise than that reported in healthy general population. Overall, the abnormal cardiovascular response to exercise in this population reveals the presence of an occult or subclinical pathology that impairs the cardiopulmonary functional reserve and reduces the efficiency of body’s oxygen delivery machinery, which could be disadvantageous during the times of increased cardiopulmonary output demands as in physiological distress or disease.

## Data Availability

Data

## Authors’ Contributions

Conceived and designed the current manuscript study: MA

Developed study protocols: WMG, MA

Collected samples and data: SZ, MD, JB, WMG, MA

Analyzed and interpreted data: SZ, MD, JB, JK, MA

Prepared and edited the manuscript: SZ, MD, JB, WMG, JK, MA

Obtained funding: MA

## Declaration/Summary conflict of interest

Authors report no conflict of interest related to this work

## Funding

This work was supported by:

1. The Flight Attendant Medical Research Institute (FAMRI) (012500WG and CIA190001 to MA).
2. California Tobacco-Related Disease Research Program (TRDRP) (T29IR0715 to MA).
3. The Department of Veterans Affairs (CXV-00125 to MA).
4. National Library of Medicine Training Grant (NIH: T15LM007442 to SZ)

The funders had no role in study design, data collection and analysis, decision to publish, or preparation of the manuscript. The statements and conclusions in this publication are those of the authors and not necessarily those of the funding agencies. The mention of commercial products, their source, or their use in connection with the material reported herein is not to be construed as an actual or implied endorsement of such products.

## Notation of prior abstract publication/presentation

Some of the results of this study have been previously reported in the form of an abstract (American Thoracic Society International Meeting 2020: Am J Respir Crit Care Med 2020;201:A7591)

## Acknowledgement

The authors would like to thank Wendy Ching, BS, Charlotte Hunt, BS, and Warren M Gold, MD, for their assistance with performing the cardiopulmonary exercise testing, and Dr. Jorge Kizer from the University of California and San Francisco VA Healthcare System Division of Cardiology for his input on the manuscript.

## Data Availability Statements

The datasets generated during and/or analyzed during the current study are available from the corresponding author on reasonable request.

## ABBREVIATION LIST

COPD: Chronic obstructive pulmonary disease
CT: Computerized tomography
DM: Density mask
EELV: End-expiratory lung volume
EFL: Expiratory flow limitation
FAMRI: Flight Attendant Medical Research Institute
FEF_25-75_: Forced expiratory flow at 25% to 75% of FVC
FEF_75_: Forced expiratory flow at 75% of FVC
FEV_1_: Forced expiratory volume in 1 second
FVC: Forced vital capacity
GLI: Global Lung Function Initiative
GOLD: Global Initiative on Obstructive Lung Disease
HU: Hounsfield unit
IC: Inspiratory capacity
LLN: Lower limit of normal
MEF: Maximum expiratory flow
PFT: Pulmonary function test
RV: Residual volume
SHS: Secondhand tobacco smoke
TLC: Total lung capacity
V_E_: Minute ventilation
V_FL_: Volume of the tidal breath that is flow-limited on expiration
Vo_2Peak_: Peak oxygen uptake
V_T_: Tidal volume
Watts_Peak_: Peak work achieved in watts
Watts-Minutes: Cumulative work achieved in watts over the entire course of exercise test

## Notes

### Competing Interest Statement

This work was supported by:
1.Flight Attendant Medical Research Institute (FAMRI) (012500WG and CIA190001 to MA).
2.California Tobacco-Related Disease Research Program (TRDRP) (T29IR0715 to MA).
3.The Department of Veterans Affairs (CXV-00125 to MA).
4.National Library of Medicine Training Grant (NIH: T15LM007442 to SZ)
The funders had no role in study design, data collection and analysis, decision to publish, or preparation of the manuscript. The statements and conclusions in this publication are those of the authors and not necessarily those of the funding agency. The mention of commercial products, their source, or their use in connection with the material reported herein is not to be construed as an actual or implied endorsement of such products.

### Funding Statement

This work was supported by:
1.Flight Attendant Medical Research Institute (FAMRI) (012500WG and CIA190001 to MA).
2.California Tobacco-Related Disease Research Program (TRDRP) (T29IR0715 to MA).
3.The Department of Veterans Affairs (CXV-00125 to MA).
4.National Library of Medicine Training Grant (NIH: T15LM007442 to SZ)

### Author Declarations

The UCSF Institutional Review Board (IRB) and the San Francisco VA Medical Center Committee on Research and Development approved study protocols. Written IRB-approved informed consent and Health Insurance Portability and Accountability Act (HIPAA) were obtained from all study participants. Subjects received monetary compensation for their participation in the study.

### Summary of Updates

This is revised to address the addition of a co-author and minor revisions in language.

## REFERENCES

1. Centers for Disease Control and Prevention. Vital Signs: Disparities in Nonsmokers’ Exposure to Secondhand Smoke - United States, 1999–2012. 2015; https://www.cdc.gov/mmwr/preview/mmwrhtml/mm6404a7.htm?s_cid=mm6404a7_w. Accessed November 28, 2018.

2. Collaborators GBDT. Smoking prevalence and attributable disease burden in 195 countries and territories, 1990-2015: a systematic analysis from the Global Burden of Disease Study 2015. Lancet. 2017;389(10082):1885–1906.

3. Tsai J, Homa DM, Gentzke AS, et al. Exposure to Secondhand Smoke Among Nonsmokers - United States, 1988-2014. MMWR Morb Mortal Wkly Rep. 2018;67(48):1342-1346.

4. Barnoya J, Glantz SA. Cardiovascular effects of secondhand smoke: nearly as large as smoking. Circulation. 2005;111(20):2684–2698.

5. Meyers DG, Neuberger JS, He J. Cardiovascular effect of bans on smoking in public places: a systematic review and meta-analysis. J Am Coll Cardiol. 2009;54(14):1249–1255.

6. Yankelevitz DF, Henschke CI, Yip R, et al. Second-hand tobacco smoke in never smokers is a significant risk factor for coronary artery calcification. JACC Cardiovasc Imaging. 2013;6(6):651–657.

7. The Health Consequences of Involuntary Exposure to Tobacco Smoke: A Report of the Surgeon General. Atlanta (GA)2006.

8. The Health Consequences of Smoking-50 Years of Progress: A Report of the Surgeon General. Atlanta (GA)2014.

9. Secondhand Smoke Exposure and Cardiovascular Effects: Making Sense of the Evidence. Washington (DC)2010.

10. Repace J. Flying the smoky skies: secondhand smoke exposure of flight attendants. Tob Control. 2004;13 Suppl 1:i8–19.

11. Benowitz NL, Bernert JT, Caraballo RS, Holiday DB, Wang J. Optimal serum cotinine levels for distinguishing cigarette smokers and nonsmokers within different racial/ethnic groups in the United States between 1999 and 2004. Am J Epidemiol. 2009;169(2):236–248.

12. Arjomandi M, Haight T, Redberg R, Gold WM. Pulmonary function abnormalities in never-smoking flight attendants exposed to secondhand tobacco smoke in the aircraft cabin. J Occup Environ Med. 2009;51(6):639–646.

13. Arjomandi M, Haight T, Redberg R, Gold WM. Pulmonary function abnormalities in never-smoking flight attendants exposed to secondhand tobacco smoke in the aircraft cabin. J Occup Environ Med. 2009;51(6):639–646.

14. Arjomandi M, Haight T, Sadeghi N, Redberg R, Gold WM. Reduced exercise tolerance and pulmonary capillary recruitment with remote secondhand smoke exposure. PLoS One. 2012;7(4):e34393.

15. Arjomandi M, Zeng S, Geerts J, et al. Lung volumes identify an at-risk group in persons with prolonged secondhand tobacco smoke exposure but without overt airflow obstruction. BMJ Open Respir Res. 2018;5(1):e000284.

16. Eisner MD, Wang Y, Haight TJ, Balmes J, Hammond SK, Tager IB. Secondhand smoke exposure, pulmonary function, and cardiovascular mortality. Ann Epidemiol. 2007;17(5):364–373.

17. Fletcher CM, Elmes PC, Fairbairn AS, Wood CH. The significance of respiratory symptoms and the diagnosis of chronic bronchitis in a working population. British medical journal. 1959;2(5147):257-266.

18. Quanjer PH, Stanojevic S, Cole TJ, et al. Multi-ethnic reference values for spirometry for the 3-95-yr age range: the global lung function 2012 equations. Eur Respir J. 2012;40(6):1324–1343.

19. Stocks J, Quanjer PH. Reference values for residual volume, functional residual capacity and total lung capacity. ATS Workshop on Lung Volume Measurements. Official Statement of The European Respiratory Society. Eur Respir J. 1995;8(3):492–506.

20. Wasserman K, Hansen J, Sue D, Stringer W, Whipp B. Principles of Exercise Testing and Interpretation. 4th edition ed: Lippincott Williams & Wilkins, Philadelphia, USA.; 2004.

21. ATS/ACCP Statement on cardiopulmonary exercise testing. Am J Respir Crit Care Med. 2003;167(2):211–277.

22. Moreno LF, Stratton HH, Newell JC, Feustel PJ. Mathematical coupling of data: correction of a common error for linear calculations. J Appl Physiol (1985). 1986;60(1):335-343.

23. Watz H. The Lungs and the Heart. American Journal of Respiratory and Critical Care Medicine. 2015;192(1):7–8.

24. Smith BM, Kawut SM, Bluemke DA, et al. Pulmonary Hyperinflation and Left Ventricular Mass. Circulation. 2013;127(14):1503–1511.

25. Cuttica MJ, Colangelo LA, Shah SJ, et al. Loss of Lung Health from Young Adulthood and Cardiac Phenotypes in Middle Age. American Journal of Respiratory and Critical Care Medicine. 2015;192(1):76–85.

26. Butler J, Schrijen F, Henriquez A, Polu J-M, Albert RK. Cause of the Raised Wedge Pressure on Exercise in Chronic Obstructive Pulmonary Disease. American Review of Respiratory Disease. 1988;138(2):350–354.

27. Tingley D, Yamamoto T, Hirose K, Keele L, Imai K. mediation: R Package for Causal Mediation Analysis. 2014. 2014;59(5):38.

28. Myers J, Kaminsky LA, Lima R, Christle JW, Ashley E, Arena R. A Reference Equation for Normal Standards for VO(2) Max: Analysis from the Fitness Registry and the Importance of Exercise National Database (FRIEND Registry). Prog Cardiovasc Dis. 2017;60(1):21–29.

29. Arjomandi M, Zeng S, Barjaktarevic I, et al. Radiographic Lung Volumes Predict Progression to COPD in Smokers with Preserved Spirometry in SPIROMICS. Eur Respir J. 2019.

30. Zeng S, Tham A, Bos B, Jin J, Giang B, Arjomandi M. Lung volume indices predict morbidity in smokers with preserved spirometry. Thorax. 2018.

31. Hendriks T, van Dijk R, Alsabaan NA, van der Harst P. Active Tobacco Smoking Impairs Cardiac Systolic Function. Scientific Reports. 2020;10(1):6608.

32. Cuttica MJ, Colangelo LA, Shah SJ, et al. Loss of Lung Health from Young Adulthood and Cardiac Phenotypes in Middle Age. American Journal of Respiratory and Critical Care Medicine. 2015;192(1):76–85.

33. Fagard R, Staessen J, Thijs L, Amery A. Prognostic significance of exercise versus resting blood pressure in hypertensive men. Hypertension. 1991;17(4):574–578.

34. Fagard RH, Pardaens K, Staessen JA, Thijs L. Prognostic value of invasive hemodynamic measurements at rest and during exercise in hypertensive men. Hypertension. 1996;28(1):31–36.

35. Mottram PM, Haluska B, Yuda S, Leano R, Marwick TH. Patients with a hypertensive response to exercise have impaired systolic function without diastolic dysfunction or left ventricular hypertrophy. J Am Coll Cardiol. 2004;43(5):848–853.

36. Takamura T, Onishi K, Sugimoto T, et al. Patients with a hypertensive response to exercise have impaired left ventricular diastolic function. Hypertens Res. 2008;31(2):257–263.

37. Filipovsky J, Ducimetiere P, Safar ME. Prognostic significance of exercise blood pressure and heart rate in middle-aged men. Hypertension. 1992;20(3):333–339.

38. Manolio TA, Burke GL, Savage PJ, Sidney S, Gardin JM, Oberman A. Exercise blood pressure response and 5-year risk of elevated blood pressure in a cohort of young adults: the CARDIA study. Am J Hypertens. 1994;7(3):234–241.

39. Tsumura K, Hayashi T, Hamada C, Endo G, Fujii S, Okada K. Blood pressure response after two-step exercise as a powerful predictor of hypertension: the Osaka Health Survey. J Hypertens. 2002;20(8):1507–1512.

40. Le VV, Mitiku T, Sungar G, Myers J, Froelicher V. The blood pressure response to dynamic exercise testing: a systematic review. Prog Cardiovasc Dis. 2008;51(2):135–160.

41. Scott JA, Coombes JS, Prins JB, Leano RL, Marwick TH, Sharman JE. Patients with type 2 diabetes have exaggerated brachial and central exercise blood pressure: relation to left ventricular relative wall thickness. Am J Hypertens. 2008;21(6):715–721.

42. Kramer CK, Leitão CB, Canani LH, Ricardo ED, Pinto LC, Gross JL. Blood pressure responses to exercise in type II diabetes mellitus patients with masked hypertension. J Hum Hypertens. 2009;23(9):620–622.

43. Allen AM, Oncken C, Hatsukami D. Women and Smoking: The Effect of Gender on the Epidemiology, Health Effects, and Cessation of Smoking. Curr Addict Rep. 2014;1(1):53–60.

44. Kamimura D, Cain LR, Mentz RJ, et al. Cigarette Smoking and Incident Heart Failure. Circulation. 2018;137(24):2572–2582.

45. Nadruz W, Claggett B, Gonçalves A, et al. Smoking and Cardiac Structure and Function in the Elderly. Circulation: Cardiovascular Imaging. 2016;9(9):e004950.

46. Rutten-van Mölken M, Roos B, Van Noord JA. An empirical comparison of the St George’s Respiratory Questionnaire (SGRQ) and the Chronic Respiratory Disease Questionnaire (CRQ) in a clinical trial setting. Thorax. 1999;54(11):995–1003.

47. Crapo RO, Morris AH, Gardner RM. Reference values for pulmonary tissue volume, membrane diffusing capacity, and pulmonary capillary blood volume. Bull Eur Physiopathol Respir. 1982;18(6):893–899.

