## Supplemental Appendix for "Remote exposure to secondhand tobacco smoke is associated with lower exercise capacity through effects on oxygen pulse, a proxy of cardiac stroke volume"

SHS Exposure Characterization

SHS exposure was characterized by a questionnaire developed by the UCSF Flight Attendant Medical Research Institute (FAMRI) Center of Excellence,^4^ and modified to acquire information on airline-related occupational history, as described previously (UCSF FAMRI SHS Questionnaire).^1,2^ Briefly, this included employer airlines, duration of employment, and flight routes with quantification of “cabin SHS exposure” as the number of years during which the crewmembers were exposed to SHS in aircraft. Other possible sources of SHS exposure were also explored by questioning participants about their non-cabin exposures in additional settings, as described previously.^5^

Pulmonary Function Testing

Lung function measurement procedures were conducted according to the American Thoracic Society (ATS) and European Respiratory Society (ERS) guidelines.^6-11^ Routine pulmonary function tests were performed in the seated position using a model Vmax 229 CareFusion (CareFusion Corp., Yorba Linda, CA) and nSpire body plethysmograph (nSpire Health Inc., Longmont, CO) as described previously.^1^ This included the spirometry measurements of flows at low lung volumes;^12^ lung volume by single breath dilution and plethysmography;^13-15^ airway resistance during panting at FRC;^16,17^ and single breath carbon monoxide diffusing capacity.^18^ Bronchodilator responsiveness was not performed. Hyperinflation and air trapping, which is inferred from an increase in FRC or RV, were quantified using the ratios of FRC or RV to TLC (FRC/TLC or RV/TLC).

Cardiopulmonary Exercise Testing

Participants performed physician-supervised, symptom-limited, progressively increasing exercise tests in the supine position on an electromagnetically braked, supine cycle ergometer (Medical Positioning Inc. Kansas City, MO). The protocol consisted of 3-minute rest, 1-minuteunloaded (freewheeling) cycling at 60 to 65 revolution per minute (rpm), followed by increasing work rate of 20 to 40 Watts at 2-minute intervals (stages) to a maximum toleration, and ending with 5-minutes of recovery. Participants were encouraged to give their best effort and were encouraged to continue exercise until a VO_2_ plateau effect on a breath-to-breath analysis of oxygen consumption was visually observed during testing; however, they were advised that they could stop voluntarily at any time they believed they could not continue.

Twelve-lead electrocardiogram (ECG), heart rate (HR), and oxyhemoglobin saturation by pulse oximetry (SpO_2_) were monitored continuously, and blood pressure (BP) (measured manually by a physician using a sphygmomanometer) was recorded every 2 minutes during the second minute of each stage. Minute ventilation (V_E_), oxygen uptake (VO_2_), and carbon dioxide output (VCO_2_) were measured breath-by-breath with an open-circuit metabolic cart (model Vmax 229, CareFusion, Yorba Linda, CA). Immediately before all tests, the gas analyzers were calibrated using reference gases of known concentrations and the ventilometer was calibrated using a 3-liter syringe (Hans Rudolph, Kansas, MO). The metabolic system was verified using four trained technicians who provided monthly exercise values as biological standards for the laboratory.

Peak exercise gas exchange variables (VO_2_, VCO_2_, and V_E_) were estimated as the last 30-second average value obtained during the highest stage of the exercise test that the participant was able to complete (Watts_Peak_), as defined by continuous cycling at the required 60 to 65 rpm during that stage for greater than 1 minute. The volumes of the flow meter, mouthpiece, and filter (70 mL x breathing frequency) were subtracted from V_E_ for the V_E_/VCO_2_ calculations. Anaerobic threshold (AT) was determined by the V-slope method.^19,20^

The participant’s perception of level of effort and exertion, breathlessness, and fatigue was documented during the second minute of every stage using the modified Borg Rating of Perceived Exertion (Borg), with the Category-Ratio Scale anchored at number 10 (CR10).^21^

4.	Eisner MD, Wang Y, Haight TJ, Balmes J, Hammond SK, Tager IB. Secondhand smoke exposure, pulmonary function, and cardiovascular mortality. *Ann Epidemiol.* 2007;17(5):364-373.

5.	van Koeverden I, Blanc PD, Bowler RP, Arjomandi M. Secondhand Tobacco Smoke and COPD Risk in Smokers: A COPDGene Study Cohort Subgroup Analysis. *COPD.* 2015;12(2):182-189.

6.	Standardization of Spirometry, 1994 Update. American Thoracic Society. *Am J Respir Crit Care Med.* 1995;152(3):1107-1136.

7.	Macintyre N, Crapo RO, Viegi G, et al. Standardisation of the single-breath determination of carbon monoxide uptake in the lung. *Eur Respir J.* 2005;26(4):720-735.

8.	Miller MR, Crapo R, Hankinson J, et al. General considerations for lung function testing. *Eur Respir J.* 2005;26(1):153-161.

9.	Miller MR, Hankinson J, Brusasco V, et al. Standardisation of spirometry. *Eur Respir J.* 2005;26(2):319-338.

10.	Pellegrino R, Viegi G, Brusasco V, et al. Interpretative strategies for lung function tests. *Eur Respir J.* 2005;26(5):948-968.

11.	Wanger J, Clausen JL, Coates A, et al. Standardisation of the measurement of lung volumes. *Eur Respir J.* 2005;26(3):511-522.

12.	Comroe Jr. JH. Pulmonary Function Tests. *Methods in Medical Research*. Chicago, Illinois: Year Book Publishers, Inc.; 1950:188.

13.	Dubois AB, Botelho SY, Bedell GN, Marshall R, Comroe JH, Jr. A rapid plethysmographic method for measuring thoracic gas volume: a comparison with a nitrogen washout method for measuring functional residual capacity in normal subjects. *J Clin Invest.* 1956;35(3):322-326.

14.	Mitchell MM, Renzetti AD, Jr. Evaluation of a single-breath method of measuring total lung capacity. *Am Rev Respir Dis.* 1968;97(4):571-580.

15.	Burns CB, Scheinhorn DJ. Evaluation of single-breath helium dilution total lung capacity in obstructive lung disease. *Am Rev Respir Dis.* 1984;130(4):580-583.

16.	Dubois AB, Botelho SY, Comroe JH, Jr. A new method for measuring airway resistance in man using a body plethysmograph: values in normal subjects and in patients with respiratory disease. *J Clin Invest.* 1956;35(3):327-335.

17.	Briscoe WA, Dubois AB. The relationship between airway resistance, airway conductance and lung volume in subjects of different age and body size. *J Clin Invest.* 1958;37(9):1279-1285.

18.	Blakemore WS, Forster RE, Morton JW, Ogilvie CM. A standardized breath holding technique for the clinical measurement of the diffusing capacity of the lung for carbon monoxide. *J Clin Invest.* 1957;36(1 Part 1):1-17.

19.	Beaver WL, Wasserman K, Whipp BJ. A new method for detecting anaerobic threshold by gas exchange. *J Appl Physiol.* 1986;60(6):2020-2027.

20.	Wasserman K, Hansen JE, Sue DY, BJ W. *Principles of Exercise Testing and Interpretation.* Philadelphia: Lea & Febiger; 1987.

21.	Borg G. *Borg's perceived exertion and pain scales.* Champaign, IL, US: Human Kinetics; 1998.

22.	Fletcher CM, Elmes PC, Fairbairn AS, Wood CH. The significance of respiratory symptoms and the diagnosis of chronic bronchitis in a working population. *British medical journal.* 1959;2(5147):257-266.

23.	Quanjer PH, Stanojevic S, Cole TJ, et al. Multi-ethnic reference values for spirometry for the 3-95-yr age range: the global lung function 2012 equations. *Eur Respir J.* 2012;40(6):1324-1343.

24.	Stocks J, Quanjer PH. Reference values for residual volume, functional residual capacity and total lung capacity. ATS Workshop on Lung Volume Measurements. Official Statement of The European Respiratory Society. *Eur Respir J.* 1995;8(3):492-506.

25.	Wasserman K, Hansen J, Sue D, Stringer W, Whipp B. *Principles of Exercise Testing and Interpretation.* 4th edition ed: Lippincott Williams & Wilkins, Philadelphia, USA.; 2004.

26.	ATS/ACCP Statement on cardiopulmonary exercise testing. *Am J Respir Crit Care Med.* 2003;167(2):211-277.

27.	Moreno LF, Stratton HH, Newell JC, Feustel PJ. Mathematical coupling of data: correction of a common error for linear calculations. *J Appl Physiol (1985).* 1986;60(1):335-343.

28.	Watz H. The Lungs and the Heart. *American Journal of Respiratory and Critical Care Medicine.* 2015;192(1):7-8.

29.	Smith BM, Kawut SM, Bluemke DA, et al. Pulmonary Hyperinflation and Left Ventricular Mass. *Circulation.* 2013;127(14):1503-1511.

30.	Cuttica MJ, Colangelo LA, Shah SJ, et al. Loss of Lung Health from Young Adulthood and Cardiac Phenotypes in Middle Age. *American Journal of Respiratory and Critical Care Medicine.* 2015;192(1):76-85.

31.	Butler J, Schrijen F, Henriquez A, Polu J-M, Albert RK. Cause of the Raised Wedge Pressure on Exercise in Chronic Obstructive Pulmonary Disease. *American Review of Respiratory Disease.* 1988;138(2):350-354.

32.	Tingley D, Yamamoto T, Hirose K, Keele L, Imai K. mediation: R Package for Causal Mediation Analysis. *2014.* 2014;59(5):38.

**SUPPLEMENTAL TABLES**

**Table S1- Associations of cardiovascular response to exercise with years of exposure to cabin SHS or air trapping.**

|  | **Independent variables** | | | | | | | | |
| --- | --- | --- | --- | --- | --- | --- | --- | --- | --- |
|  | **Cabin SHS exposure** | | | **RV/TLC** | | | **FRC/TLC** | | |
| **Dependent variables** | **N** | **PE±SEM**  **(95% CI)** | **P value** | **N** | **PE±SEM**  **(95% CI)** | **P value** | **N** | **PE±SEM**  **(95% CI)** | **P value** |
| **Peaks** |  |  |  |  |  |  |  |  |  |
| O_2_-Pulse_Peak_ | **241** | **-0.032±0.015**  **(-0.062 to -0.001)** | **0.040** | 235 | -0.058±0.032  (-0.121 to 0.005) | 0.069 | 220 | -0.005±0.025  (-0.055 to 0.044) | 0.829 |
| HR_Peak_ | 241 | -0.009±0.124  (-0.253 to 0.235) | 0.941 | **235** | **-0.521±0.257**  **(-1.028 to -0.014)** | **0.043** | **220** | **-0.397±0.195**  **(-0.782 to -0.012)** | **0.043** |
| SBP_Peak_ | 223 | -0.070±0.151  (-0.368 to 0.228) | 0.642 | 220 | 0.259±0.304  (-0.340 to 0.858) | 0.394 | 213 | 0.064±0.236  (-0.402 to 0.530) | 0.786 |
| DBP_Peak_ | 223 | -0.109±0.078  (-0.262 to 0.044) | 0.162 | 220 | 0.069±0.159  (-0.244 to 0.382) | 0.663 | 213 | 0.009±0.123  (-0.233 to 0.251) | 0.942 |
| **Slopes** |  |  |  |  |  |  |  |  |  |
| O_2_-Pulse_Slope_ | 234 | 0.001±0.001  (-0.001 to 0.002) | 0.345 | 229 | 0.001±0.002  (-0.002 to 0.004) | 0.475 | 218 | 0.001±0.001  (-0.002 to 0.003) | 0.605 |
| HR_Slope_ | 234 | 0.016±0.007  (0.001 to 0.030) | 0.034 | 229 | -0.001±0.015  (-0.031 to 0.028) | 0.929 | 218 | -0.012±0.011  (-0.034 to 0.010) | 0.289 |
| SBP_Slope_ | 224 | 0.004±0.010  (-0.016 to 0.023) | 0.724 | 221 | 0.024±0.020  (-0.015 to 0.064) | 0.228 | 214 | -0.005±0.016  (-0.036 to 0.026) | 0.758 |
| DBP_Slope_ | 224 | 0.001±0.005  (-0.009 to 0.012) | 0.814 | 221 | 0.002±0.011  (-0.019 to 0.023) | 0.820 | 214 | 0.005±0.008  (-0.011 to 0.022) | 0.508 |

Footnote: The association between each of the cardiovascular response to exercise (dependent variable) and years of exposure to cabin SHS or air trapping (independent variable) was individually assessed by linear regression modeling with adjustment for age, sex, height, BMI, and corresponding baseline values (for slope variables). Statistical significance was determined by a P value<0.05. Abbreviations- SHS: secondhand smoke, O_2_-Pulse: oxygen uptake per heartbeat; HR: heart rate; SBP: systolic blood pressure; DBP: diastolic blood pressure.

**Table S2- Mediation analysis of effect of air trapping on exercise capacity through mediators.**

|  | **Mediation analysis of effect of FRC/TLC to** | | | | | | | | |
| --- | --- | --- | --- | --- | --- | --- | --- | --- | --- |
|  | **Watts_Peak_** | | | **Work_Total_** | | | **VO_2Peak_** | | |
| **Mediators** | **N** | **% mediated**  **(95% CI)** | **P value** | **N** | **% mediated**  **(95% CI)** | **P value** | **N** | **% mediated**  **(95% CI)** | **P value** |
| **Cardiovascular outputs** | | | | | | | | | |
| **Peaks** |  |  |  |  |  |  |  |  |  |
| **O_2_-Pulse_Peak_** | 220 | 28.1(-705 to 396.6) | 0.628 | 219 | 22.2(-328.5 to 414.5) | 0.658 |  | N/A * |  |
| **HR_Peak_** | 220 | 51.2(-648.3 to 515.8) | 0.396 | 219 | 56.5(-814 to 660.2) | 0.364 | 220 | 53.0(-416.6 to 693.5) | 0.230 |
| **SBP_Peak_** | 213 | 1.4(-171.2 to 172.6) | 0.920 | 213 | 1.4(-212.2 to 210.7) | 0.932 | 213 | 2.1(-111.4 to 130) | 0.848 |
| **DBP_Peak_** | 213 | 0.002(-49.3 to 73.3) | 0.999 | 213 | 0.1(-58.3 to 38.1) | 0.960 | 213 | 0.03(-29.3 to 27) | 0.962 |
| **Slopes** | | | | | | | | | |
| **O_2_-Pulse_Slope_** | 218 | 12.9(-217.9 to 193.5) | 0.540 | 218 | 10.4(-166.8 to 188.6) | 0.560 |  | N/A * |  |
| **HR_Slope_** | 218 | 16.3(-370 to 336.6) | 0.570 | 218 | 10.3(-256.1 to 224.6) | 0.590 | 218 | 8.2(-138.9 to 148.5) | 0.496 |
| **SBP_Slope_** | 214 | 3.1(-160.8 to 229.7) | 0.864 | 214 | 1.4(-91.4 to 140.6) | 0.852 | 214 | 2.2(-82.1 to 110.7) | 0.808 |
| **DBP_Slope_** | 214 | 9.8(-176.3 to 190.4) | 0.544 | 214 | 6.5(-108.8 to 145.6) | 0.570 | 214 | 9.0(-92.7 to 110.1) | 0.520 |
| **Pulmonary outputs** | | | | | | | | | |
| **RR_Peak_** | 214 | 41.3(-446 to 482.9) | 0.302 | 214 | 50.0(-540.2 to 635.1) | 0.348 | 214 | 63.2(-544 to 619.5) | 0.222 |
| **VE_Peak_** | 220 | 73.3(-991 to 1198.2) | 0.368 | 219 | 76.0(-920.1 to 1072.6) | 0.340 | 220 | 81.1(-812.4 to 781.2) | 0.202 |
| **VT_Peak_** | 218 | 21.1(-289.9 to 261.4) | 0.512 | 218 | 16.8(-279.5 to 251.4) | 0.546 | 218 | 13.6(-140 to 209.2) | 0.506 |

Footnote: To see if the association of air trapping (independent variable) with exercise capacity (dependent variable) was mediated through the cardiovascular or pulmonary outputs, mediation analyses were performed with adjustment for age, sex, height, and BMI and inclusion of one mediator at a time. Statistical significance was determined by a P value<0.05. * The associations of VO_2Peak_ with O_2_-Pulse_Peak_ and O_2_-Pulse_Slope_ were not computed due to existence of “mathematical coupling” between VO_2_ and O_2_-Pulse, which depends on VO_2_ for its calculation (O_2_-Pulse=VO_2_/HR).

Abbreviations- VO_2Peak_: peak oxygen uptake; SHS: secondhand smoke; O_2_-Pulse_Peak_: oxygen uptake per heartbeat at peak exercise; HR: heart rate; SBP: systolic blood pressure; DBP: diastolic blood pressure; RR_Peak_: peak respiratory rate; VE_Peak_: peak minute ventilation value; VT_Peak_: peak tidal volume; FRC: functional residual capacity; TLC: total lung capacity.
